# Intersecting single-cell transcriptomics and genome-wide association studies identifies crucial cell populations and candidate genes for atherosclerosis

**DOI:** 10.1101/2021.11.23.21266487

**Authors:** Lotte Slenders, Lennart P. L. Landsmeer, Kai Cui, Marie A.C. Depuydt, Maarten Verwer, Joost Mekke, Nathalie Timmerman, Noortje A.M. van den Dungen, Johan Kuiper, Menno P.J. Winther, Koen H.M. Prange, Wei Feng Ma, Clint L. Miller, Redouane Aherrahrou, Mete Civelek, Gert J. de Borst, Dominique P.V. de Kleijn, Folkert W. Asselbergs, Hester M. den Ruijter, Arjan Boltjes, Gerard Pasterkamp, Sander W. van der Laan, Michal Mokry

## Abstract

**Background:** Genome-wide association studies have discovered hundreds of common genetic variants for atherosclerotic disease and cardiovascular risk factors. The translation of susceptibility loci into biological mechanisms and targets for drug discovery remains challenging. Intersecting genetic and gene expression data has led to the identification of candidate genes. However, previously studied tissues are often non-diseased and heterogeneous in cell composition, hindering accurate candidate prioritization. Therefore, we analyzed single-cell transcriptomics from atherosclerotic plaques for cell-type-specific expression to identify atherosclerosis-associated candidate gene-cell pairs.

**Methods and Results:** To identify disease-associated genes, we applied gene-based analyses using GWAS summary statistics from 46 atherosclerotic and cardiovascular disease, risk factors, and other traits. We then intersected these candidates with scRNA-seq data to identify genes specific for individual cell (sub)populations in atherosclerotic plaques. The coronary artery disease loci demonstrated a prominent signal in plaque smooth muscle cells (*SKI*, *KANK2*, *SORT1*) p-adj. = 0.0012, and endothelial cells (*SLC44A1*, *ATP2B1*) p-adj. = 0.0011. Further sub clustering revealed genes in risk loci for coronary calcification specifically enriched in a synthetic smooth muscle cell population. Finally, we used liver-derived scRNA-seq data and showed hepatocyte-specific enrichment of genes involved in serum lipid levels.

**Conclusion:** We discovered novel gene-cell pairs, on top of known pairs, pointing to new biological mechanisms of atherosclerotic disease. We highlight that loci associated with coronary artery disease reveal prominent association levels in mainly plaque smooth muscle and endothelial cell populations. We present an intuitive single-cell transcriptomics-driven workflow rooted in human large-scale genetic studies to identify putative candidate genes and affected cells associated with cardiovascular traits. Collectively, our workflow allows for the identification of cell-specific targets relevant for atherosclerosis and can be universally applied to other complex genetic diseases and traits.

**Translational perspective:** GWAS identified a large number of genomic loci associated with atherosclerotic disease. The translation of these results into drug development and faster diagnostics remains challenging. With our approach, we cross-reference the GWAS findings for atherosclerotic disease with scRNA-seq data of disease-relevant tissue and bring the GWAS findings closer to the functional and mechanistic studies.

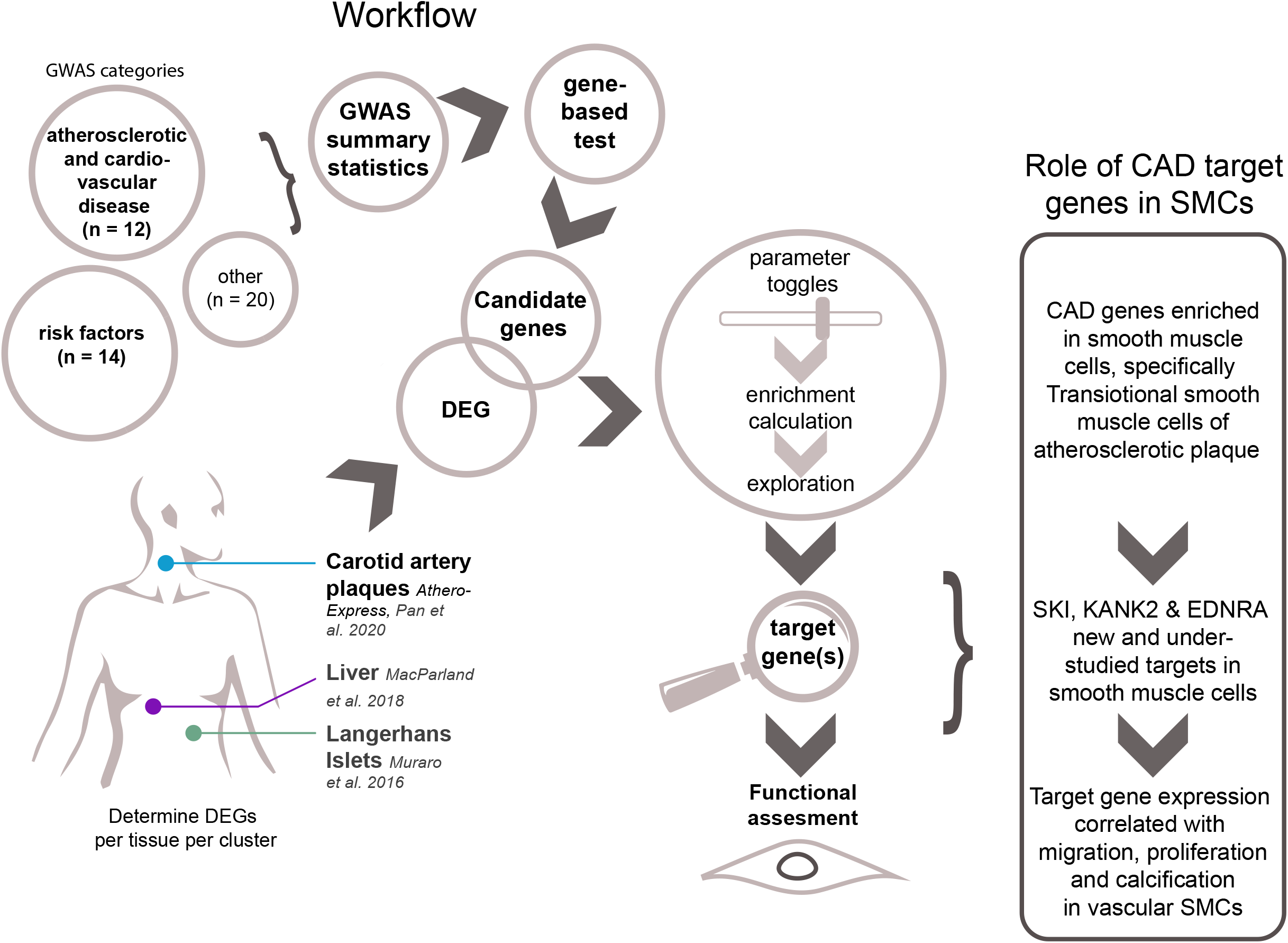

## Introduction

Since the first publication in 2005 on age-related macular degeneration^1^, genome-wide association studies (GWAS) soared, and to date the GWAS Catalog^2^ registered over 5,000 publications covering thousands of genetic associations. These efforts also identified hundreds of common genetic risk loci for cardiovascular diseases and traits, including coronary artery disease (CAD)^3^, ischemic stroke subtypes^4^, coronary artery calcification^5^, and carotid intima and media thickness^6^. Despite tremendous socio-economic and medical progress, cardiovascular disease remains the leading cause of death^7^, highlighting the need for new biomarkers and pharmaceutical targets. Previous studies convincingly show that GWAS offer a solid foundation for innovative drug development^8^ as they agnostically identify genetic relationships with diseases and point to potential causal mechanisms^9^.

Identifying the causal genes and underlying pathological mechanisms that drive the signal in genetic risk loci is not straightforward. One possible approach to narrow down the lists of candidate genes is to utilize gene expression data, by identifying expression quantitative trait loci (eQTLs) that colocalize with genetic risk loci with expression changes in relevant tissues^10^. Alternatively, transcriptome-wide association studies (TWAS) estimate genetic correlation between tissue-specific gene expression and complex traits to predict causal relations between traits^11^. However, these powerful methods take advantage of whole-tissues collected in large biobanks and the variable cell composition confounds the resulting signal. Because tissues are composed of many cell types and tissue expression is averaged across all those cells, the signal from less prevalent but important cell populations is obscured. In order to test potential therapeutic targets, one requires suitable functional assays. This in turn requires an idea of the function and cellular expression of targets, which is complicated by tissue heterogeneity. Single-cell transcriptomics enables measurement of cell-specific expression, cell-to-cell expression variability, transcriptional noise, or temporal dynamics^12^. Intersecting GWAS data with human transcriptomic single-cell data would increase the resolution both in terms of candidate gene and more importantly, it would identify the cell type most relevant for the given disease.

We hypothesised that genetic drivers for atherosclerotic disease can be found in plaque tissue. We aimed to project GWAS signals for 46 traits directly into a cell-type specific expression using single-cell RNA sequencing (scRNA-seq) data derived from atherosclerotic plaques. To this end, we devised a workflow that utilized gene-based testing to identify candidate genes from GWAS summary statistics and calculated an enrichment score for each candidate list using expression profiles from scRNA-seq driven cell populations. This approach allows us to find the enrichment in expression patterns that point to specific, and potentially causal, cell populations. We report that loci associated with coronary artery disease reveal prominent association in mainly plaque smooth muscle cells and endothelial cells.

## Methods

### Single-cell sequencing: carotid artery plaques

Atherosclerotic lesions were collected from 12 female and 26 male patients undergoing a carotid endarterectomy procedure. All pathological tissue was included in the Athero-Express Biobank Study biobank (AE, www.atheroexpress.nl) at the University Medical Centre Utrecht (UMCU)^13^. Plaque processing for single-cell sequencing is described elsewhere^14^. After sequencing, data was processed in an R 3.5 and 4.0 environment^15^ using Seurat version 3.2.2^16^. Mitochondrial genes were excluded and doublets were omitted by gating for unique reads per cell (between 400 - 10 000) and total reads per cell (between 700 – 30 000). Data was corrected for sequencing batches using the function SCTransform. Clusters were created with 30 principal components at resolution 0.8. Multiple iterations of clustering were used to determine optimal clustering parameters. Population identities were assigned by evaluating gene expression per individual cell clusters. Sub-populations of SMC populations were determined by isolating the SMC and EC populations from the complete population of cells mentioned above and re-assigned clusters. Clusters were created with 10 principal components at resolution 1.4. New subpopulation identities were assigned by evaluating gene expression per individual cell clusters. The newly defined SMC subpopulation identities were projected onto the SMC populations in the complete scRNA-seq dataset of atherosclerotic lesions for downstream processing.

### GWAS summary statistics

GWASs summary statistics were leveraged from publicly available datasets. Mining these datasets uncovered a plethora of challenges. Not all GWAS summary statistics are available for download, can be requested by the authors, and/or only provide the lead SNPs. Other issues included ambiguous reports of sample size, genome build or missing information such as standard errors and effect sizes. We were able to partly resolve these issues by using a custom pipeline, which automates tasks such as genome build liftover and SNP alignment and provides a uniform file format (**Supplemental Figure 2A**). This significantly decreased workload and improved downstream applications.

GWAS summary statistics were selected by means of relevance, “latest and the largest” and publicly available, easily accessible and/or open-source data was given priority. Incomplete summary statistics were omitted during this phase (**Supplemental Figure 2A**). A custom Python^17^ pipeline (see **Data availability**) was used to pre-process GWAS summary statistics and primary quality control. This primary control step was used to uniformly transform GWAS summary statistics because of a lack of a standardized format. In short, our custom pipeline was built to automate effect/other allele alignment with 1000 Genomes phase 3^18^ release with a maximum frequency distance of 25%, maximum MAF of 45% for ambivalent variants and lifting genomic positions to hg19 where needed. In preparation for GWAS alignment, dbSNP153 (GCF1405) was referenced to translate MEGASTROKE rs-ids to genetic positions and to augment AF, ASD, NICM and TAGC with allele frequency information. Standard errors for HF and NICM were recovered as follows:

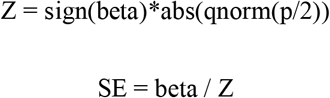

Where beta is the effect size, p is the p-value of association which is quantile normalized using the qnorm-function in R. SE is the standard error calculated by dividing the beta by Z. Effect sizes were calculated from odds-ratios as beta=log(OR) for BIP, Insomnia, IBD and MDD before entering our custom pipeline.

The final selection was grouped into one of 3 categories: atherosclerotic disease and other cardiovascular disease (12), risk factors (14) and other (20). Data was further processed with MAGMA^19^ (version 1.07) for genome-wide analysis, annotation and characterisation of significant hits via the FUMA^20^ web platform (version 1.3.6, accessed June 2020, https://fuma.ctglab.nl/). We applied genome-wide significance (p < 5×10^−8^) and linkage disequilibrium (r^2^> 0.05) filtering for clumping of independent loci within 1000 kb of the lead variant, and included only variants with minor allele frequency (MAF) > 1%. For SVS, logOnset, EvrSmk and SVD no SNP associations below the genome wide significance threshold we found. Therefore lead variant discovery was performed with a relaxed threshold of 5×10^−6^.

### Selecting cell population specific genes

DEGs were selected via the Wilcoxon sum rank test. A gene was considered a DEG if 1) 10% of cells within a cluster expressed the gene with log_2_ fold change of at least 0.25; 2) the gene passed the Bonferroni adjusted significance threshold of p < 0.05 for this test. DEGs were determined via a “one cluster vs one cluster” and “one cluster vs remaining clusters”^5^ (**Supplemental Figure 2B**). This method results in genes specific for a cluster compared to every other single cluster and genes specific for a cluster compared to the whole single cell reference set.

### Gene score calculation

Fully processed summary statistics and cell population specific DEG were used for calculating gene scores. The background gene set (n = 20112 genes) was defined as all the genes that are reported and annotated by MAGMA, regardless of their respective p value or Z score with duplicates excluded. We calculated the random gene score per cell-population by overlapping the genes in this background set with the cell-population specific DEG. For every anaysis, the background genes are the same per GWAS. Z-score threshold for the p values from MAGMA was set at 3 and we included a maximum of 150 candidate genes meeting this threshold (**Supplemental Figure 1B**). If there were more than 150 genes meeting the threshold, we only used the first 150. Calculations were made as follows: 1) Per cell population, determine the overlap of the candidate genes of a GWAS with the cell population specific DEG. 2) Score each of these genes according to average cell population expression: assign one point for expression between 1 and 2, and two points for expression > 2 and 0 points for expression < 1. Only genes that are cell population specific receive a score. Assign the points to the genes from the random candidate gene list in the same way (**Supplemental Figure 2Cii**). 3) Compare the scores from the candidate genes with the scores of the random genes via one tailed Wilcoxon Rank sum test in R and adjust the results per GWAS for multiple testing using the Benjamini-Hochberg method. Adjusted p-values smaller than 0.1 (10% false discovery rate) were considered significant (**Supplemental Figure 2Ciii**). Enrichment was defined as the fold change of the mean of the gene scores divided over the mean of the gene scores in the random set. All scripts are available on GitHub (see **data availability**).

## Author contributions

L.S. drafted the manuscript and designed the figures. M.V., J.M. and N.T. collected clinical data. N.v.d.D. executed human plaque processing and general maintenance of the biobank. G.J.d.B. performed carotid endarterectomy procedures. G.P. and G.J.d.B. participated in the design of the Athero-Express Biobank Study. M.A.C.D. executed the human plaque processing, FACS and flow cytometry. L.S., M.M., and S.W.v.d.L. participated in conceptualisation, data collection and data interpretation. L.P.L.L. and S.W.v.d.L. designed and executed workflow for raw GWAS summary statistics processing. L.S. performed data analysis on scRNA-seq data and gene score calculations. W.F.M and C.M. designed and integrated the plaque scRNA-seq into plaqview. R.A. and M.C. provided data for functional validation. S.W.v.d.L., F.W.A., M.M., J.K., M.W. and G.P. participated in the conceptualization, funding and supervision of the scRNA-seq experiments. All authors provided feedback on the research, analyses and manuscript.

## Results

### Single-cell RNA-seq of atherosclerotic plaques reveals 18 cell populations

We have expanded our previously published^14^ scRNA-seq datasets of human atherosclerotic plaque cells in a cohort which now includes 38 patients and identified 18 cell populations amongst 6,191 cells. These expanded datasets allowed us to map the genetic loci not only to main cell types (e.g., smooth muscle cells (SMCs)) but also to more specific cell subtypes (e.g., synthetic, contractile or transitioning SMCs) - increasing resolution compared to the previous dataset. By combining known and recently published sets of markers we identified clusters of lymphocytes, macrophages, mast cells, endothelial cells (ECs), and SMCs (**Figure 1A**). This data is available for interactive exploration via the online tool Plaqview^21^ accessible on www.plaqview.com. Notably, the expression of selected, widely studied GWAS targets for coronary artery disease (*EDN1*, *HDAC9*, *NOS3*, *VEGFA*, *FN1*, *APOE*, *APOB*, *SMAD3*, *PCSK9* and *LDLR*) clearly demonstrates cell-specific patterns (**Figure 1B**). This underscores the need for a systematic analysis using large-scale GWAS summary statistics and motivated us to devise an intuitive two-stage approach.

**Figure 1.**
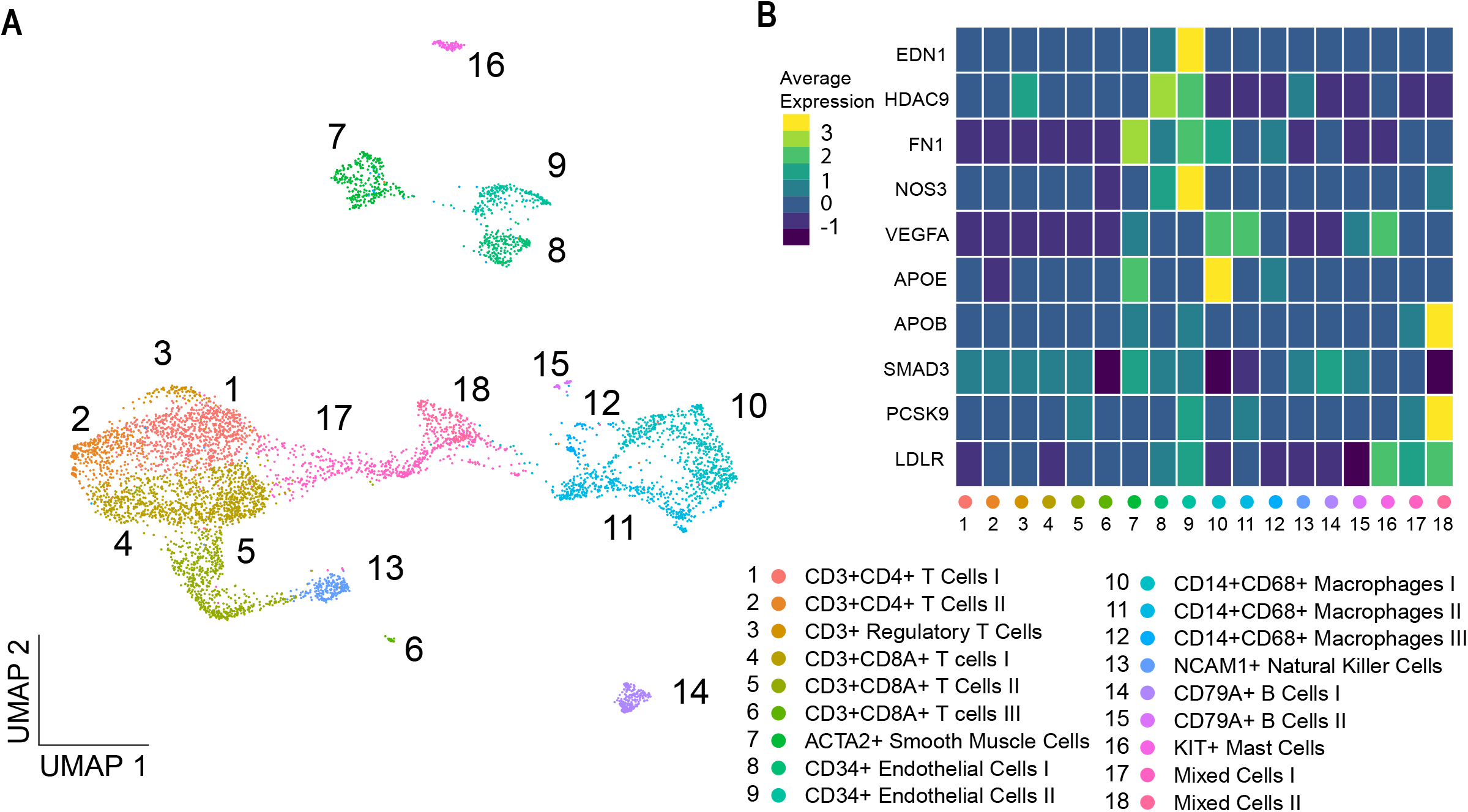
Finding suitable therapeutic targets from GWAS with scRNA-seq. **A)** UMAP visualisation of 6,191 cells divided into 18 cell populations derived from 38 human carotid artery plaques. **B)** Heatmap of frequently studied CAD GWAS hits: *EDN1*, *HDAC9*, *NOS3*, *VEGFA*, *FN1*, *APOE*, *APOB*, *SMAD3*, *PCSK9* and *LDLR* in atherosclerotic tissue shown in cells depicted in cell populations shown in **A**.

### Gene-based analyses of atherosclerotic diseases and cell population specific genes

First, we aimed to identify trait-associated genes using GWAS summary statistics by aggregating per-variant statistics and to calculate an empirically derived p-value for a given gene. Thus, gene-trait associations would be agnostically derived and solely based on genetic signals. To do this we collected and partitioned GWAS summary statistics for 46 traits across 3 categories: “atherosclerotic disease” (4), “cardiometabolic traits” (22) and “other” (20) (**Supplemental Table 1, Supplemental Figure 1A**). From these GWASs, 40 were performed on population with European ancestry, and the remaining 6 had mixed or other ancestries. Next, to identify genes associated with individual traits we performed gene-based testing using MAGMA^19^ per individual GWAS. Genes mapped by gene-based testing are referred to as *candidate genes* (**Supplemental Figure 2A**, **Supplemental Data**). Second, we aimed for an unbiased selection of genes that would best represent each cell population in a given dataset (**Supplemental Figure 2B**). Per tissue, these genes were selected through differential gene expression and include genes that are overexpressed in specific cell populations compared to all other cells (one vs. all) or compared to any individual cell cluster (one vs. one)^14^. This approach eliminates ubiquitously expressed genes and genes with low expression without the need of a predefined list (**Supplemental Figure 4**) and identifies a similar number of cell-specific genes in each population. Cell population specific genes will be further referred to as *differentially expressed genes* (DEGs). For cell populations in the atherosclerotic plaque, the average number of DEG was 461, with the lowest 307 for B Cells II and highest 678 DEGs for T Cells III.

### Plaque cell populations show specific enrichment of GWAS candidate genes

To test our hypothesis that the genetic drivers for atherosclerotic disease can be found in plaque tissue, we applied our workflow on cardiovascular candidate genes and the DEG sets from individual cell types present in the atherosclerotic plaque (**Supplemental Figure 3A, C, Supplemental Figure 4, Supplemental Table 2, Supplemental Data**). The signals we observed were independent of the total number of DEGs in each cell population (**Supplemental Figure 3C**), and did not show a bias towards GWAS sample size (**Supplemental Figure 1C**) or trait heritability (**Supplemental Figure 1D**). We found cell-specific enrichment for different GWAS in the atherosclerotic disease category, which was mainly observed in ECs and SMCs. Stroke candidate genes were enriched in EC populations, Regulatory T Cells and B Cells II. (**Figure 3A**).

**Figure 2.**
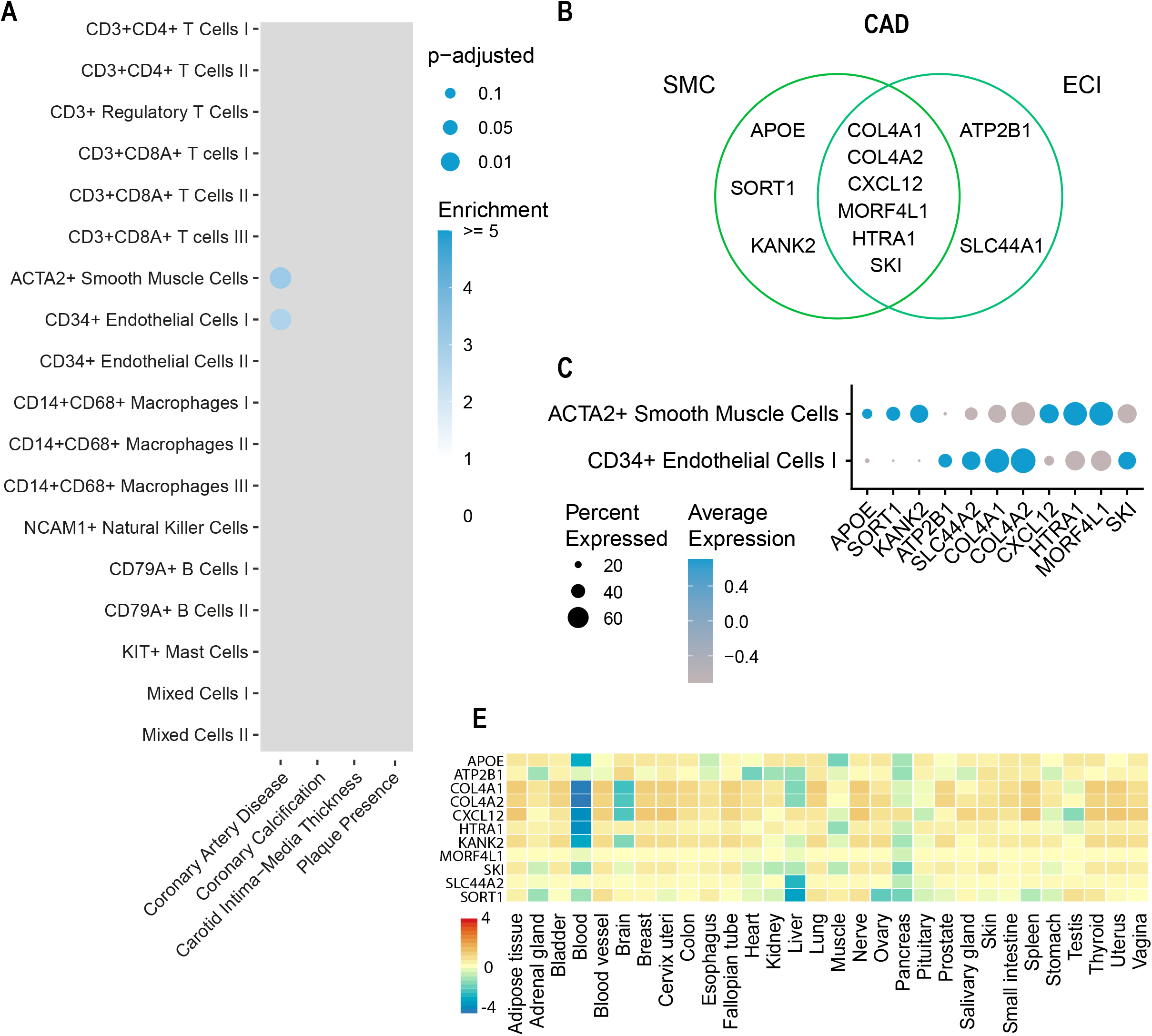
GWAS candidate genes show cell-specific enrichment in atherosclerotic plaque cell populations. **A**) Plot representing overlaps of GWAS candidate genes on cardiovascular disease with genes specifically expressed in cell populations from human atherosclerotic plaque. Colour intensity indicates enrichment over random data and size indicates adjusted p value (Benjamini-Hochberg). **B**) Genes for CAD underlying the overlaps in SMC and ECI populations. Only significant overlaps are plotted. **C**) Dotplot depicting expression of genes from **B** in the respective cell populations. **D**) Average of normalized expression (zero mean across samples) of genes from CAD in SMCs from **A** in 30 tissues from GTEx.

**Figure 3.**
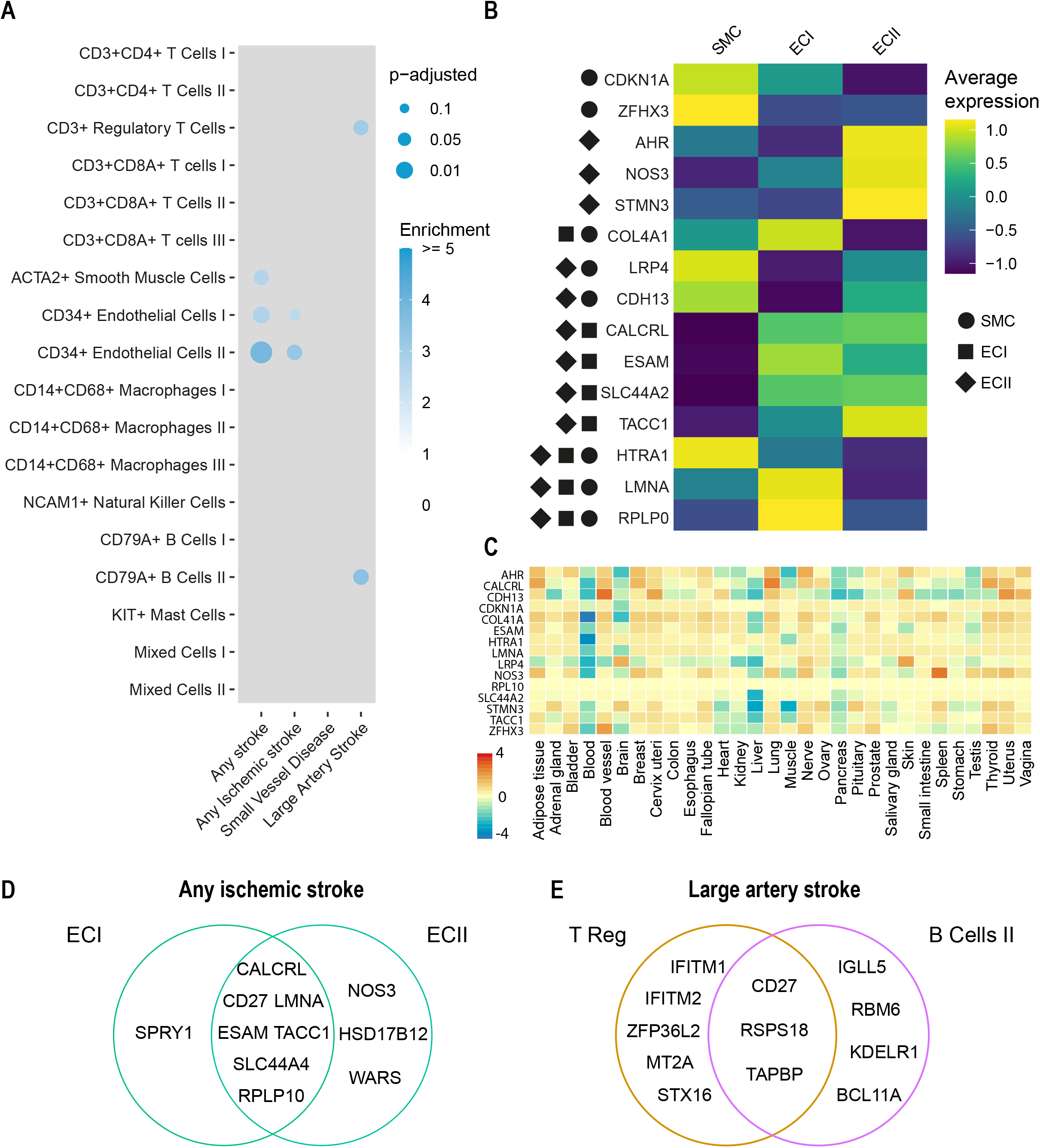
GWASs related to stroke show substrate in Endothelial cells. **A**) Plot representing overlaps of GWAS candidate genes on stroke with genes specifically expressed in cell populations from human atherosclerotic plaque. Colour intensity indicates enrichment over random data and size indicates adjusted p value (Benjamini-Hochberg). **B**) Heatmap depicting expression of genes from any stroke from **A** in the respective cell populations. Only significant overlaps are plotted, symbols represent the cell populations with the significant overlaps. **C**) Average of normalized expression (zero mean across samples) of genes from any stroke in from **B** in 30 tissues from GTEx. **D**) Genes for any ischemic stroke underlying the overlaps in ECI and ECII populations. Only significant overlaps are plotted. **E**) Genes for large artery stroke underlying the overlaps in T Reg and B cells II populations. Only significant overlaps are plotted.

### Smooth muscle cells are predominantly associated with coronary artery disease genes

By highlighting the overlap between plaque specific DEGs and candidate genes for CAD, we observed a significant enrichment in SMCs (p-adj. = 0.0012, 3.1 fold enrichment) and ECI (p-adj. = 0.0011, 2.7 fold enrichment) populations (**Figure 2A**). All p values and fold enrichment values are listed in **Supplemental Table 2**. This uncovered well studied GWAS loci such as *APOE, COL4A1* and *COL4A2*. The remainder of the overlap is driven by *KANK2* and *SORT1* in SMCs, *SLC44A1* and *ATP2B1* in ECI (**Figure 2B**). *KANK2* belongs to the KANK protein family which plays a role in cytoskeletal function. *SKI* expression was present in both cell populations (**Figure 2C**). This proto-oncogene inhibits TGF-beta signalling^22^. Interestingly, while using the single cell transcriptomics enabled us to pinpoint specific cell (sub)populations, these potential targets exhibited ubiquitous expression in bulk transcriptomics data from GTEx tissue samples (**Figure 2D**). Notably, CD14+CD68+ Macrophages III showed an enrichment of 4.3 for CAD with genes *APOE*, *SKI* and *HNRNPUL1* but did not meet the significance threshold (p-adj. = 0.19).

To verify our results we used an independent scRNA-seq dataset of carotid artery plaques by Pan *et al.*^23^ (**Supplemental Figure 3B, D**). The cell population labels broadly correspond between datasets (**Supplemental Figure 3E**). The patterns for enrichment are similar between datasets (**Supplemental Figure 3A, B**). We found that CAD, coronary artery calcification and plaque presence candidate genes were enriched in the Fibrochondrocyte cell population (**Figure 3B**), which corresponds to ACTA2+ SMCs cells in our dataset. For CAD, the enriched was driven by genes *CDKN2A*, *COL4A1*, *COL4A2*, *APOC1*, *APOE*, and *CXCL12*. Similar to the genes found in our datasets in SMCs and ECs (**Supplemental Table 2**).

### Endothelial cells are enriched for ischemic stroke-associated genes

Atherosclerotic disease is one of the major culprits underlying stroke, with carotid plaques being the main driver of large artery stroke. We subsequently investigated overlap of GWAS-associated genes for stroke subtypes any stroke (AS) and any ischemic stroke (IS) and found they were predominantly enriched in ECI (AS: p-adj. = 0.011, 2.7 fold enrichment; IS: p = 0.077, 2.5 fold enrichment) and ECII (AS: p-adj = 1.9×10^−5^, 3.9 fold enrichment; IS: p-adj = 0.023, 3.2 fold enrichment) subtypes (**Figure 3A**). Overlapping genes between AS and cell populations include *ESAM*, *LMNA* and *SLC44A2*. (**Figure 3B**) Soluble ESAM (endothelial cell adhesion molecule) levels have been correlated to myocardial infarction and heart failure^24^. *Lmna* deficient mouse models develop cardiovascular disease at an accelerated pace due to premature ageing^25^. Of note, expression of these genes was not specific to vascular derived tissue of GTEx when using bulk transcriptomics data, suggesting that our findings are cell population and tissue specific (**Figure 3C**). No significant enrichment is found for IS in SMCs, but the cell population specific signal in ECI and ECII is largely different from AS (**Figure 3D**).

Large Artery Stroke (LAS) is a subtype of IS. Strikingly, we see enrichment in CD3+ Regulatory T Cells and CD79A+ B Cells II (plasma cells) (p-adj = 0.026 and p-adj = 0.021 resp.) populations for LAS (**Figure 3E**). Suggesting the involvement of the adaptive immune system on the development of the LAS phenotype.

### Transitional SMCs enriched for atherosclerotic disease candidate genes

Plaque ACTA2+ Smooth muscle cells can be further divided into three sub-populations with a distinct phenotype; Synthetic Smooth Muscle Cells, Contractile Smooth Muscle Cells and a smaller cell population consisting of SMCs with an intermediate phenotype (Transitional Smooth Muscle Cells) (**Figure 4A, B**, **Supplemental Figure 5A, Supplemental Table 2**). To explore if we can narrow down the signal from SMCs into a specific SMC subtype, we subjected this subset of plaque cells to our workflow (**Supplemental Figure 5B, C**). This revealed that the majority of the signal can be found in Transitional SMCs. This population is predominantly enriched for CAD (p-adj. = 0.026 2.4 fold enrichment), carotid intima-media thickness (cIMT) (p-adj. = 0.027, 2.4 fold enrichment) and plaque presence (p-adj. = 0.081, 2.3 fold enrichment) (**Figure 4C**). Intima-media thickness is a commonly used tool to measure atherosclerosis^26^. Signal for LAS was present in Transitional and Synthetic SMCs with unique hits for each cell population (**Figure 4D**).

**Figure 4.**
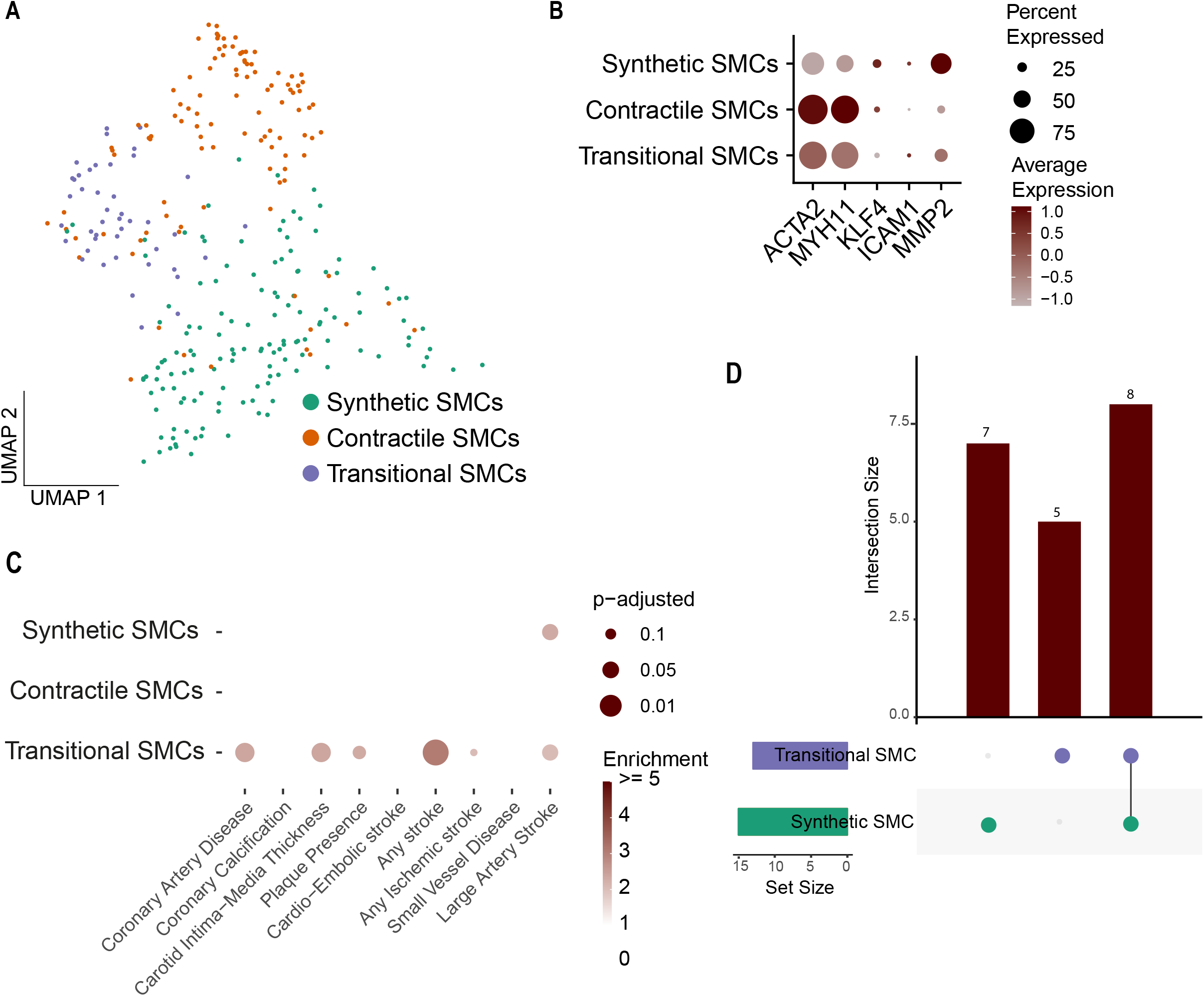
Analysis of 3 sub-populations of atherosclerotic Smooth Muscle Cells. **A**) UMAP visualisation of 3 sub cell populations of SMCs in atherosclerotic plaque. **B**) Dotplot depicting expression of synthetic and contractile SMC markers in the 3 sub-populations. **C**) Plot representing overlaps of GWAS candidate genes on cardiovascular disease and cardiometabolic traits with genes specifically expressed in the subset of SMCs from human atherosclerotic plaque. Colour intensity indicates enrichment over random data and size indicates adjusted p value (Benjamini-Hochberg) **D**) Plot depicting the number of unique and intersecting candidate genes from Large artery stroke enriched in Transitional SMCs and Synthetic SMCs.

To test the robustness of these results we have repeated the analysis with the other public dataset (Pan *et al.)*. For CAD, the overlap between Transitional SMCs and corresponding “Fibrochondrocytes” in Pan et al. (**Supplemental Figure 5DE**) showed significant enrichment and again identified a largely overlapping set of commonly studied GWAS loci *COL4A1*, *COL4A2*, *APOC1*, *APOE*, and *CXCL12*.

### Functional assessment of smooth muscle cell associated candidates

To demonstrate the potential role of newly identified gene-cell pairs in atherosclerosis-relevant process we selected 3 genes enriched in the SMC population of plaques. *SKI*, *KANK2* and *EDNRA* are CAD candidate genes and currently understudied in relation to atherosclerotic disease in this cell population. Making them possible interesting targets for further functional studies. To investigate the role of these genes in SMCs, we functionally characterized SMCs from the ascending aortas in 151 healthy and diverse donors for cellular migration, proliferation and calcification^27^. *KANK2* gene expression is correlated with proliferation under different stimuli (p-adj. < 0.1), and was negatively correlated with calcification in quiescent cells (Calcification Osteo: p-adj = 0.0058; Calcification (Pi): p-adj 0.069) (**Figure 5A**, **Supplemental Table 5**). Under all stimuli, the response was positive, except upon TGFβ1 exposure. SKI expression positively correlates with calcification in proliferative cells (p-adj = 0.069) but has a negative response towards TGFβ1 induced proliferation (p-adj = 0.069, R -0.21) (**Figure 5A**, **Supplemental Table 5**). *EDNRA* was negatively correlated with proliferation response to IL1β (p-adj. = 0.038) and calcification (p-adj = 0.062) in non-proliferative cells and showed a positive correlation in proliferative cells for migration (p-adj = 0.069) (**Figure 5A**, **Supplemental Table 5**).

**Figure 5.**
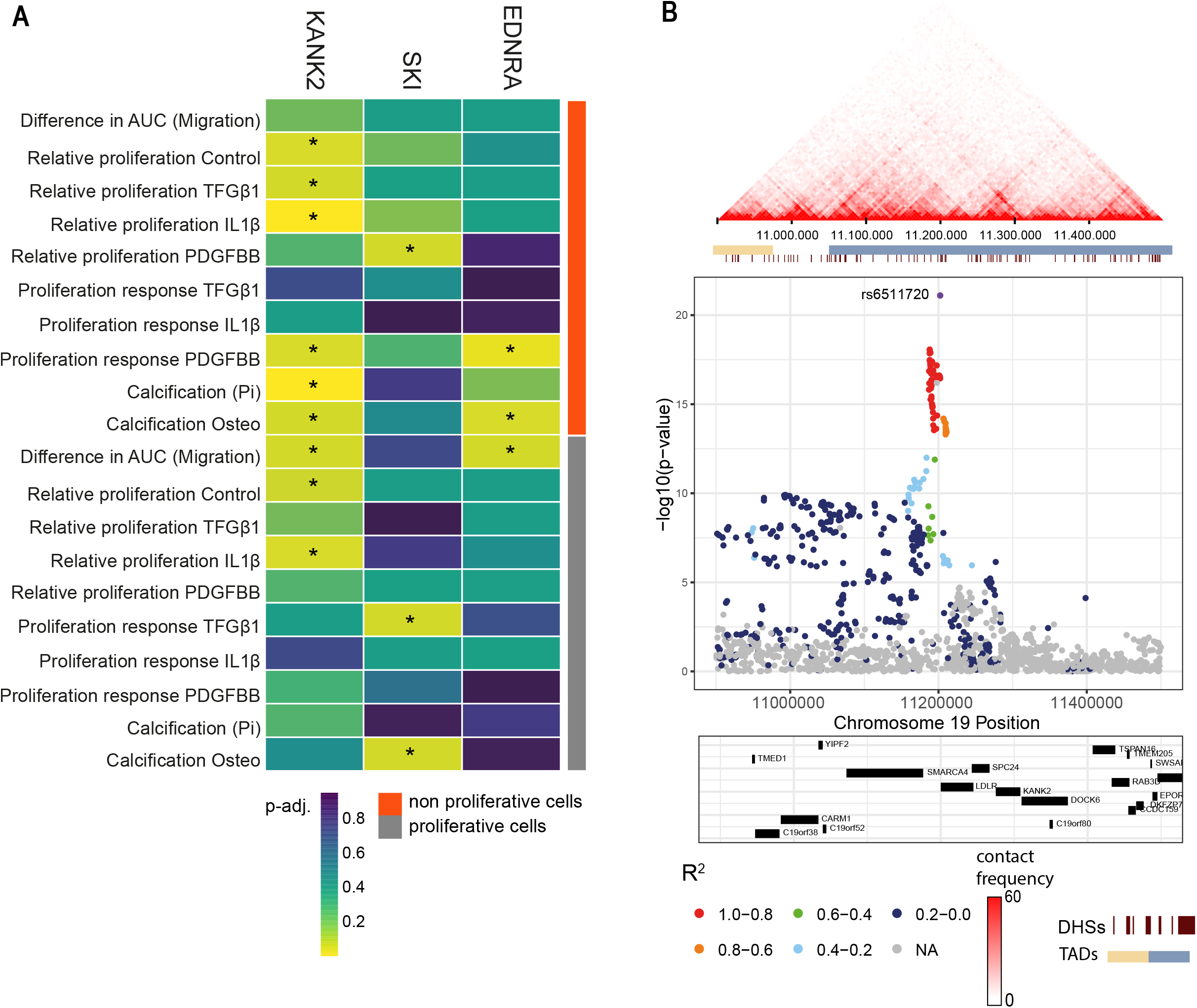
SMC associated candidates influence migration and proliferation *in vitro.* **A**) Correlation of *KANK2*, *SKIi* and *EDNRA* expression non-proliferative and proliferative cells. **B**) Plot depicting the Hi-C interactions around the main SNP associated with KANK2 (top) and regional association plot (bottom) in Hi-C data derived from IMR90 fibroblasts. AUC: Area under curve; Pi : Inorganic Phosphate; DHS: DNaseI hypersensitivity sites; TAD: Topologically associating domain.

### Lipid related risk factors enriched in liver hepatocyte populations

Causal mechanisms for atherosclerosis have been linked to lipid metabolic processes active in the liver and blood glucose levels. This offered us the opportunity to assess the robustness of our approach across tissues and diseases. We acquired published scRNA-seq datasets of human liver cells^28^ and human pancreatic Langerhans islet cells^29^ to identify possible cell- and disease-specific targets.

Since one of the major liver functions is lipid metabolism, we expected strong signals for candidate genes involved in triglycerides levels. In line with our expectations, these GWASs show indeed the strongest signal in hepatocytes 3,4 and 5 driven by several apolipoproteins (**Supplemental Figure 6A, C, Supplemental Table 2**).

Langerhans islet cells overall showed weak enrichments for all atherosclerosis related GWASs (**Supplemental Figure 5B, D**). Signal for CAD in endothelial and mesenchymal populations overlap with hits in comparable cell populations in plaque (**Supplemental Table 2**). We could detect significant overlap of type 2 diabetes associated genes with Beta cells (p-adj. = 0.001) driven by *INS*, *WFS1* and *FTO* amongst others (**Supplemental Table 2**).

Altogether, we show that candidate genes for atherosclerotic disease are significantly enriched in EC and SMC populations of atherosclerotic plaque, notably in transitional SMCs. Stroke related genes were enriched in ECs and lipid loci were found enriched in liver hepatocytes. Pancreatic beta cells were enriched for BMI candidate genes. Interestingly, only limited signals were observed in the cells of the adaptive immune system suggesting the genetic component for atherosclerosis is rooted in the structural cells of the vessel. We selected 3 candidate genes enriched in SMCs for functional testing in cells of ascending aortas for cellular calcification, proliferation and migration that showed that *KANK2, SKI* and *EDNRA* expression correlated with calcification, migration and proliferation in vascular SMCs.

## Discussion

Post-GWAS analyses aiming to identify candidate genes for translation into clinical care are not straightforward. The thousands of variants associated with complex traits, including cardiovascular diseases, have made it abundantly clear that many genes contribute to disease risk and progression. For instance, GWAS for coronary artery disease have identified 163 loci linking to hundreds of candidate genes^3^. Further efforts like chromatin interaction mapping linked almost 300 additional gene targets for atherosclerotic disease via distal chromatin interactions^14^. This adds another layer to candidate gene prioritization. Selecting causal GWAS loci for functional testing can be deduced with vast knowledge of the trait or disease, but this remains speculative. There are numerous ways to accelerate the path to translational research. The intersection of loci found amongst different GWAS related to the same trait or disease can highlight some of the primary genes involved but, the cellular resolution is still lacking^4, 30^. Overlapping GWAS hits with pathways or within gene networks offers more context to study the genes. However, this is highly dependent on knowledge of the respective pathways and networks, and locating the relevant key players in the network is not indisputable^31^. Colocalization of tissue-specific eQTLs^10, 32, 33^ with GWAS data provides another alternative that can be used to estimate if there are overlapping causal variants in both datasets. For example. a recent study on type 2 diabetes leveraged publicly available eQTL data and a separate pancreatic dataset to find targets for glucose- and insulin-related trait loci^10^. In concordance to our data, trait associated genes are expressed in pancreatic islet tissue. However cell population specific signals are lacking and eQTL signals can be confounded by variability in tissue cell composition. Currently, efforts are being made to study eQTLs in a single-cell space, with single-cell quantification of gene expression in disease relavant tissue. With this method it is possible to detangle the effect of SNPs on the single cell and patient specific level and contruct personalized gene regulatory networks ^12, 34^.

Another straightforward approach to identify the causative genes is overlapping the expression profiles of different tissues and organs to GWAS candidate genes. Utilizing bulk RNA sequencing data applicable to the GWAS helps to find targets that are directly acting in the affected tissue. This powerful approach has one major drawback: the data is composed of multiple cell populations and this signal is difficult to deconvolute into individual cell populations. It is especially limiting when looking for cells that are present in almost all tissues – like endothelial cells or smooth muscle cells.. We show here that these bulk RNA tissues approaches can be misleading due to lack of specific expression patterns for relevant GWAS loci in specific tissue, like vascular wall tissue (**Figure 2E**). On the other hand, the overlap with single cell transcriptomics can detect significant overlaps in distinct cell types such as ECs.

Here we present a possible approach to the prioritization problem. We projected GWAS loci directly into single-cell transcriptomics datasets derived from disease-relevant tissue. This allowed us to identify both known and novel gene targets together with their associated cell populations. Notably, the CAD-associated gene *SKI* is significantly enriched in SMC and EC populations (**Figure 2A**). *SKI* inhibits TGF-beta signalling^22^, which plays a major role in atherosclerotic disease progression by controlling processes such as cell proliferation and matrix formation. Consequently, *SKI* increases lesion size in LDR-/- mice^35^. Another identified gene - *KANK2* plays a role in cytoskeletal function by reducing the cell’s ability to migrate by activating *TALIN*^36^. This gene is specifically expressed in the SMC-population of plaques. Possible impairment of cell mobility by *KANK2* could lead to inefficient SMC migration towards the cap resulting in an unstable plaque phenotype. Interestingly, our functional analysis of *KANK2* showed a positive association between gene expression and cell migration in quiescent and proliferative cells of SMCs from ascending aortas (R = 0.15 and 0.2 resp.) (**Supplemental table 5**) – suggesting more complex mechanisms. Follow-up experiments creating gene-deficient primary cell lines derived from atherosclerotic plaques to study the molecular mechanistic of those genes can provide more information of its role in atherosclerosis^37^. Notably, the CAD-associated locus in the vicinity of *KANK2* also contains *LDLR* gene (**Figure 5B** bottom). Even though LDLR involvement in atherosclerosis is widely accepted, we were able to show that *KANK2* is another strong candidate in this region as it also shows a strong correlation with SMC proliferation (**Figure 5A**). This also demonstrates that our approach can identify genes that are located more further from the lead SNP but are positioned within same broader regulatory domain (**Figure 5B**) - as based on Hi-C data from IMR90 fibroblast cells^38^ (**Figure 5B** top).

Currently 32 stroke loci have been implicated by GWAS studies^4^. From the new main stroke loci found by Malik *et al.,* only *SLC44A2* was significantly enriched in scRNAseq populations. However, we identified other potential targets in SMCs, ECs and in B cells and regulatory T Cell populations (**Figure 3A**). One of the targets, *ESAM*, was not yet directly linked to atherosclerosis, but known to enhance vascular permeability in selective tissues^39^ and could therefore contribute to the influx of cholesterol and inflammatory cells in the arteries contributing to the stroke phenotype. Another target, Gs-coupled receptor calcitonin receptor–like receptor (*CALCRL*) is expressed in both ECI and ECII populations. Activation of this receptor leads to an anti-inflammatory response in endothelial cells. Subsequently *CALCRL* deficient mice present with more advanced atherosclerotic lesions^40^. A recent study by Örd *et al.* researched the enrichment of CAD, ischemic stroke and seven other cardiometabolic GWAS signals in scATAC-seq peaks of carotid artery plaques. Similarly, they report enrichment of CAD and stroke loci in SMCs and ECs and total cholesterol loci in macrophages^41^.

Remarkable is the absence of significant enrichment in the cells of the adaptive immune system of the plaque for CAD. Out of six T cell populations, four exhibited the enrichment of 0.00 and CD3+CD8A+ T Cells II and II showed enrichment of 1.3 and 0.37 respectively, indicating that there is no or very little overlap between CAD candidate genes and cell population-specific DEGs. This suggests that the genetic component of atherosclerotic disease is not rooted in the adaptive immune system but rather in the structural cells of the vessel and influences the integrity of the vascular wall or plaque stability.

Our workflow can be generalized to other single-cell transcriptomics datasets and complex traits. We applied it to scRNAseq-data from tissues with expected causal involvement in atherosclerotic disease: human liver cells and human pancreatic Langerhans Islet cells. GWASs studying lipid levels were associated with hepatocyte subsets but in particular subsets Hep3, Hep4 and Hep5 (**Supplemental Figure 6A**). It suggests that these risk factors for atherosclerosis have a stronger biological foundation in lipid metabolism in the liver-rather than local lipid metabolism in the plaque. Langerhans Islet cells show less prominent signals for our selected GWAS, suggesting that the genetic mechanisms involved in atherosclerosis do not have a local component in the pancreas (**Supplemental Figure 6B**).

Little to no signal can be observed for GWASs that have no relation to the cells composing the atherosclerotic plaque, such as height, cancers and behavioural related traits such as bipolar disorder and neuroticism (**Supplemental Figure 3A**). Yet, these GWASs have ample candidate genes and considerable GWAS sample size - suggesting the specificity of our approach. (**Supplemental Figure 1B, C, Supplemental Table 1**). Similarly, no signal for BMI is found in plaque cell populations, intuitively confirming that the mechanisms underlying obesity cannot be accounted for in plaque tissue^42^.

This research, however, is subject to several limitations. In our workflow, a bias may be introduced by low-powered GWAS, or the definition of a gene, e.g. ±50kb around the transcription start and end sites, but it is not affected by the tissue expression. Only in our second step, we introduce a prioritization step for tissue expression and cell populations. In highly heterogeneous tissues, such as atherosclerotic plaque, multiple genes can be shared amongst the same class of cell populations (e.g. all endothelial cells), whilst they are lacking in others. To resolve this problem and to keep such genes as cell type-specific, we implemented an approach that can unambiguously uncover cell-specific genes without the need of a predefined list of ubiquitously expressed genes. Although this method is inclusive, the balance between the sufficient number of differentially expressed genes and their specificity can be adapted depending on the scRNA-seq dataset in question. The depth and quality of the scRNAsequencing is a another limiting factor. The amount and kind of RNA that can be detected and favors the most abundant genes with highest mRNA counts per cell. As a result certain gene classes, like transcription factors or signaling molecules are more difficult to detect. At the same time, cell types like neutrophils and foam cells in atherosclerotic plaque are difficult to capture by single cell transcriptomics and might be underrepresented or completely missed. Digestion protocols, tissue quality and sample handling also affect the quality and/or cell composition of scRNA-seq data.

In conclusion, we systematically projected GWAS summary statistics for 46 traits directly into single-cell transcriptomic profiles across 18 distinct cell populations derived from plaques. We devised a two-step workflow that 1) leverages large-scale human genetic data to prioritize trait-associated genes, and 2) calculates a cell-type specific enrichment score. We uncovered 11 potential cell population specific targets for CAD and provided functional evidence for *KANK2*, *SKI* and *EDNRA* in smooth muscle cells. Additionally, we verified the robustness and transportability of our approach by confirming the tissue- and trait-specific enrichment of circulating lipid genes in liver and glycemic disorder genes in pancreatic tissue. The principal strength of our framework is the cell-specific resolution it offers for disease associated genes mapped to GWAS loci, thus putting the emphasis on prioritization of candidates (instead of adding to an ever growing candidate-list) and accelerating the path to functional testing.

## Data availability

Custom scripts used for this publication are available on GitHub https://github.com/CirculatoryHealth/gwas2single. Raw summary statistics can be accessed through their original publications (**Supplemental Table 4**). Quality controlled and/or FUMA-MAGMA processed summary statistics are available for download at Zenodo (https://zenodo.org/record/4823040). Raw human carotid artery plaque scRNAseq data can be accessed via https://dataverse.nl/dataverse/atheroexpress. The plaque scRNAseq data after quality control, i.e. counts, scores, etc., used in this paper can be found at https://doi.org/10.34894/TYHGEF. Other data supplements can be found in the **Supplementary Data section**.

## Data Availability

Custom scripts used for this publication are available on GitHub https://github.com/CirculatoryHealth/gwas2single. Raw summary statistics can be accessed through their original publications (Supplemental Table 4). Quality controlled and/or FUMA-MAGMA processed summary statistics are available for download at Zenodo (https://zenodo.org/record/4823040). Raw human carotid artery plaque scRNAseq data can be accessed via https://dataverse.nl/dataverse/atheroexpress. The plaque scRNAseq data after quality control, i.e. counts, scores, etc., used in this paper can be found at https://doi.org/10.34894/TYHGEF. Other data supplements can be found in the Supplementary Data section.

https://github.com/CirculatoryHealth/gwas2single

https://dataverse.nl/dataverse/atheroexpress

## Acknowledgements

The Genotype-Tissue Expression (GTEx) Project was supported by the Common Fund of the Office of the Director of the National Institutes of Health, and by NCI, NHGRI, NHLBI, NIDA, NIMH, and NINDS. The data used for the analyses described in this manuscript were obtained from: FUMA GTEx Portal on 12-07-2020.

We thank Sonya A. MacParland and Mauro J. Muraro for providing their scRNA-seq data on human liver and pancreatic islet cells at our request.

We are thankful for the support of the ERA-CVD program ‘druggable-MI-targets’ (grant number: 01KL1802), the EU H2020 TO_AITION (grant number: 848146) and the Leducq Fondation ‘PlaqOmics’ (grantnumber: 18CVD02).

## Appendices

## Supplementary files

### Supplementary methods

#### PlaqView

To facilitate the sharing and exploration of our dataset and uncovering of potential therapeutic candidates, we integrated our plaque data into PlaqView^21^: www.plaqview.com. PlaqView is a standalone, interactive, and reproducible web-based tool to explore single-cell RNA-sequencing data of cardiovascular tissue material. Within PlaqView, users can quickly query gene expression, cell trajectories, and explore drug targets within the context of the single-cell data. On PlaqView, researchers can explore our dataset, download high-quality graphical outputs without the need for prior coding experience. PlaqView is built on R and Shiny, and its source code is publicly available at https://github.com/MillerLab-CPHG/PlaqView.

#### Estimation of SNP-heritability and genetic correlation

We estimated the SNP-heritability due to genotyped and imputed SNPs (□phenotypic variance casually explained by common SNPs) of 46 atherosclerotic, cardiometabolic, and other traits using high-definition likelihood (HDL) and LD score regression (LDSC). HDL was performed using Rpackage “HDL” v1.3.9 (https://github.com/zhenin/HDL) and LDSC was performed using LDSC software v1.0.1 (https://github.com/bulik/ldsc). In LDSC analysis, GWAS summary statistics were harmonized by the *munge sumstats* procedure. In HDL analysis, we estimated the SNP-based narrow-sense heritability and those with SNPs with less than 90% overlap with the HDL reference panel (UKB_imputed_SVD_eigen99_extraction) were removed.

#### Single cell sequencing: plaque, liver and pancreas

Sequencing data from human carotid artery plaques^23^, liver^28^ and pancreas^29^ can be accessed through their original publications (**Supplemental Table 4**). Prior to downstream processing, reads from liver and pancreas were filtered for UGDH-AS1, PGM2P2, KCNQ1OT1, MALAT1, PGM5P2, MAB21L3, EEF1A1, and ERCC spike-ins. Original cell population identities were preserved and no additional filtering on cells and genes was performed in all cases.

#### Functional assay of smooth muscle cells

Vascular smooth muscle cells (SMCs) were isolated from aortic explants of anonymous donors (n=136) through the UCLA heart transplant program as described previously^27^. Cells from additional 15 donors were purchased from Lonza and PromoCell. They were maintained in Smooth Muscle Cell Basal Medium (SmBM, CC-3181, Lonza) supplemented with Smooth Muscle Medium-2 SingleQuots Kit (SmGM-2, CC-4149, Lonza) (complete media). They were cultured in complete media (containing 5% FBS) until 90% confluence. Media was then switched to either serum-free media to mimic the quiescent state of SMCs in healthy arteries or continued to culture in complete media to mimic the proliferative state of SMCs in atherosclerotic arteries for 24 hours. Total RNA was extracted with Qiagen miRNEasy kit. Sequencing libraries were prepared with the Illumina TruSeq Stranded mRNA Library Prep Kit and were sequenced to ∼100 million read depth with 150 bp paired-end reads at the Psomogen sequencing facility. The reads with low average Phred scores (<20) were trimmed using Trim Galore. They were then mapped to the hg38 version of the human reference genome using the STAR Aligner in two-pass mode to increase the mapping efficiency and sensitivity. Only the uniquely mapped read pairs were retained. Gene expression was quantified by calculating the transcripts per million (TPM) for each gene using RNA-SeQC based on GENCODE v32 transcript annotations. Proliferation, migration, and calcification were also measured in these cells. The amount of calcification was quantified in media containing high inorganic phosphate or osteogenic stimuli. These two media formulations recapitulate different aspects of arterial calcification that occur in advanced stages of atherosclerosis. The proliferation was quantified in control media and media containing platelet-derived growth factor β1 (TGF-β1), or interleukin-1β (IL-1β). Finally, the amount of migration was quantified to a PDGF-BB gradient over 24 hours using a modified Boyden chamber assay. Overall, 12 quantitative phenotypes related to calcification, proliferation, and migration were performed. Correlation between gene expression and phenotypes were calculated using Pearson Correlation Coefficient in R.

### Supplementary table list

Supplemental Table 1

Supplemental Table 2 (access via Zenodo, see data availability)

Supplemental Table 3 (access via Zenodo, see data availability)

Supplemental Table 4

Supplemental Table 5

**Supplemental Figure 1.**
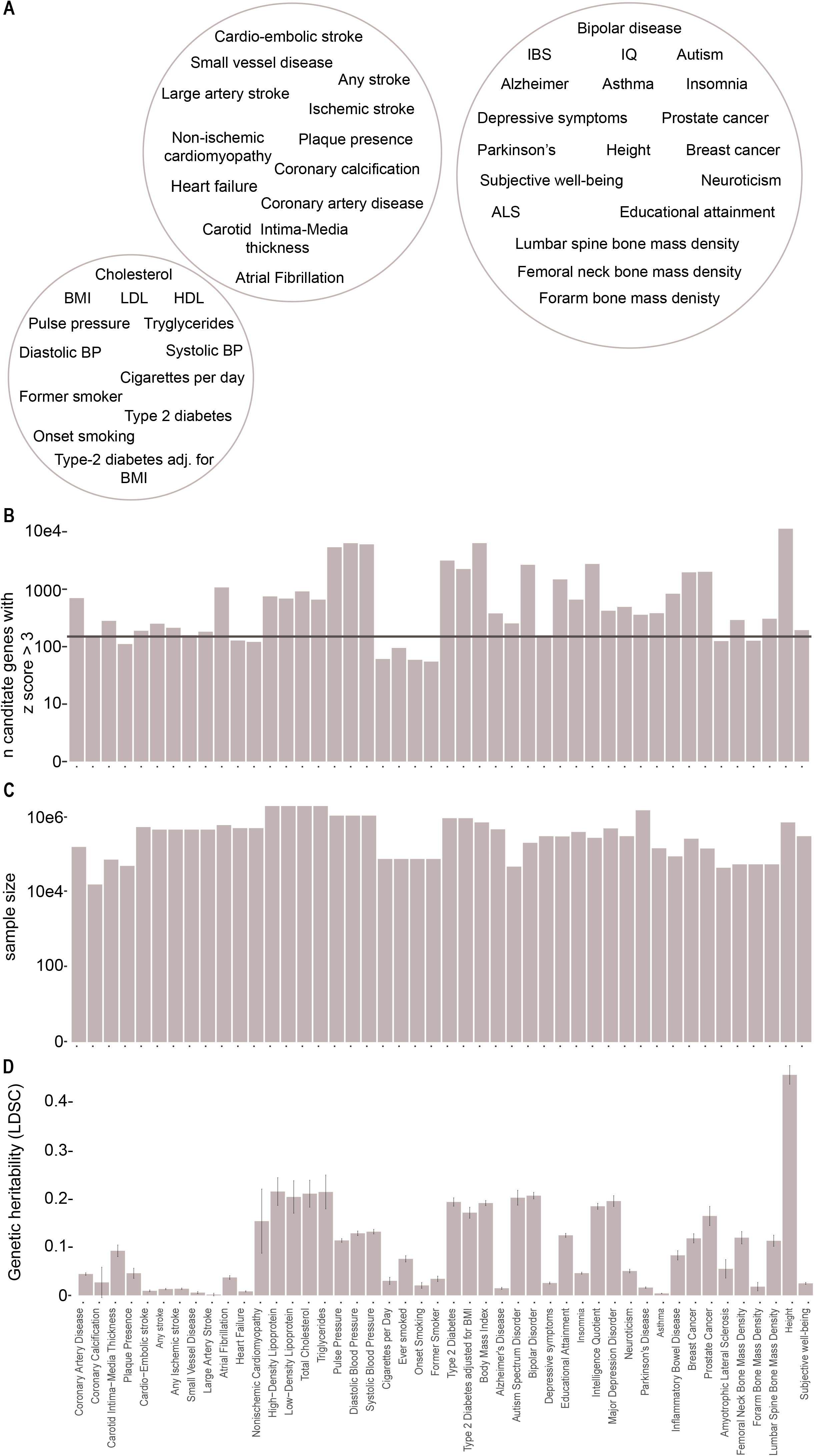
Overview of the 46 selected GWAS summary statistics. **A**) Visual representation of 46 GWAS across 3 categories: atherosclerotic disease, cardiometabolic traits, and others. **B**) Barplot depicting the n candidate genes per GWAS with Z score cutoff 3. Horizontal line indicates the threshold of max candidate genes (150) included in the gene score calculations. **C**) Barplot depicting the sample size per GWAS. **D**) Genetic heritability as calculated using LD Score Regression. LDSC: Linkage Disequilibrium Score Regression.

**Supplemental Figure 2.**
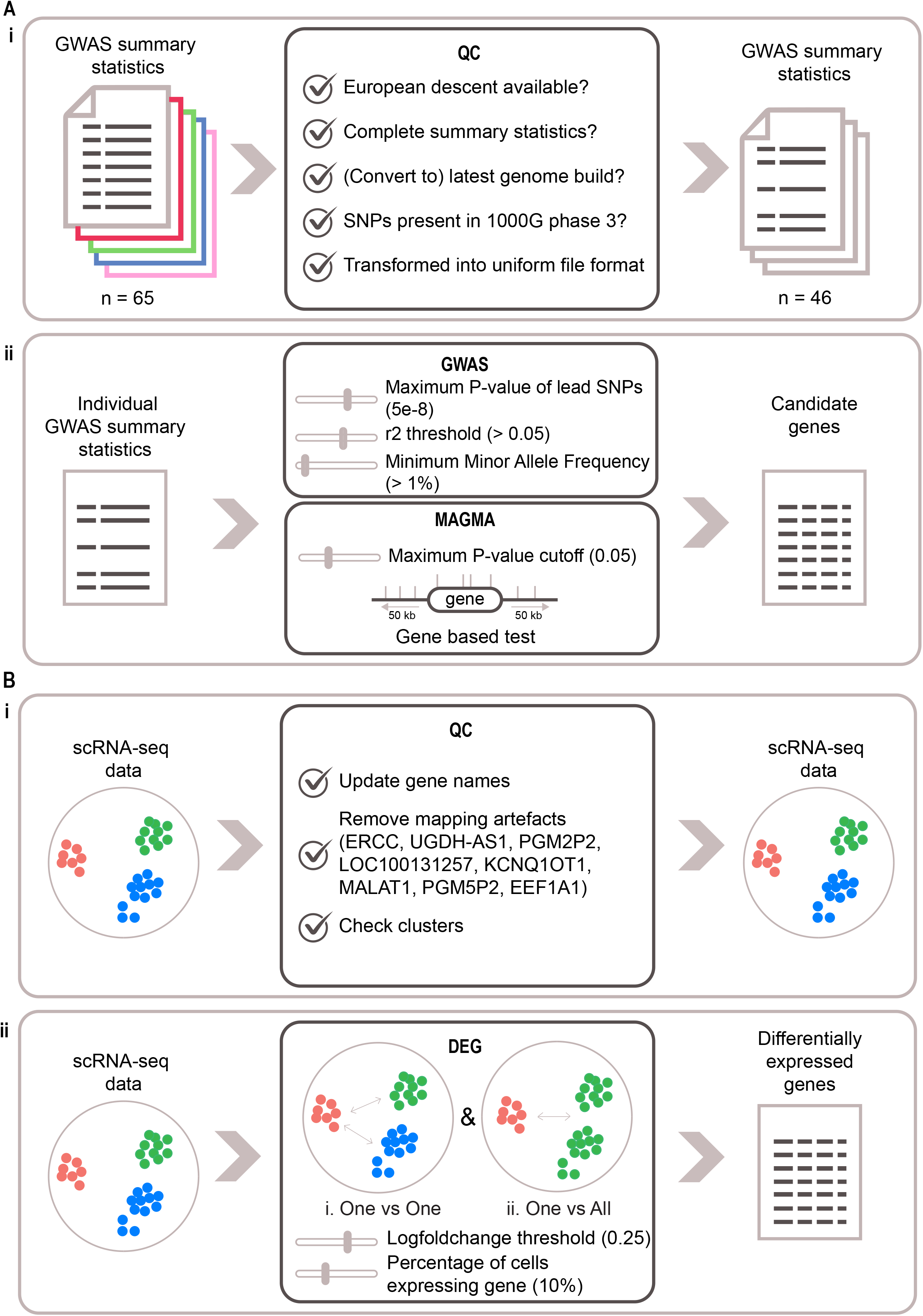
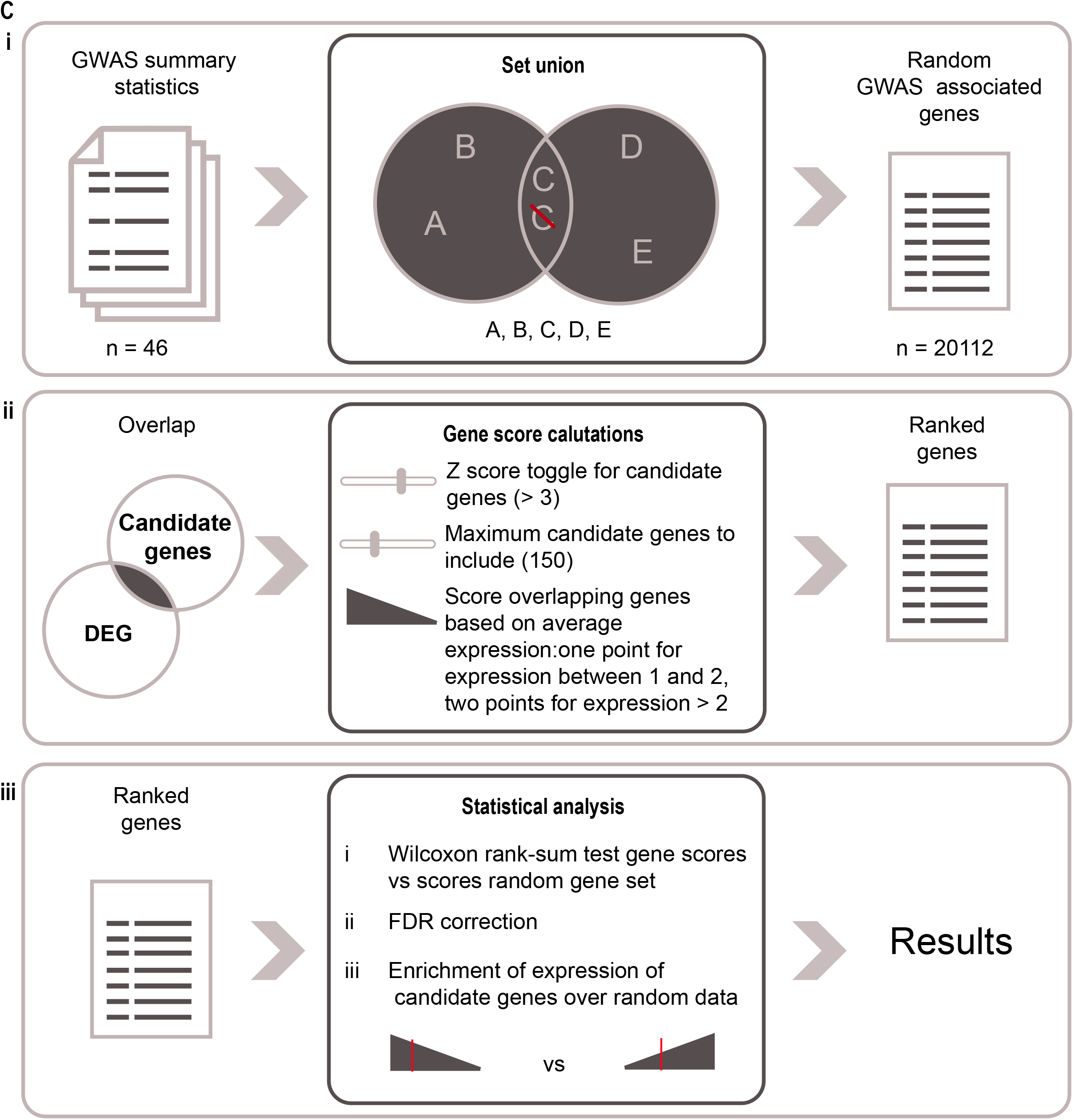
Schematic overview of *in-silico* workflow. **A**) GWAS summary statistic processing. i) GWAS summary statistics undergo quality control and are transformed into a uniform file format. ii) Individual GWAS summary statistics are processed followed by gene based testing by MAGMA, resulting in a list of associated genes per GWAS. **B**) Yielding scRNA-seq cell population specific genes. i) scRNA-seq data undergo quality control. ii) Differentially expressed genes are acquired via a “one vs. one” or “one vs. all” method. **C**) Gene score calculations. i) Generate a random GWAS dataset from all other GWASs and subject them to step i. ii) Where DEGs and candidate genes overlap, the size of the overlap can be adjusted. Remaining genes will be scored and ranked according to average expression in the scRNA-seq data, and the scores for the random data. iii) Enrichment over random genes is calculated. Results have resolution on a single gene basis.

**Supplemental Figure 3.**
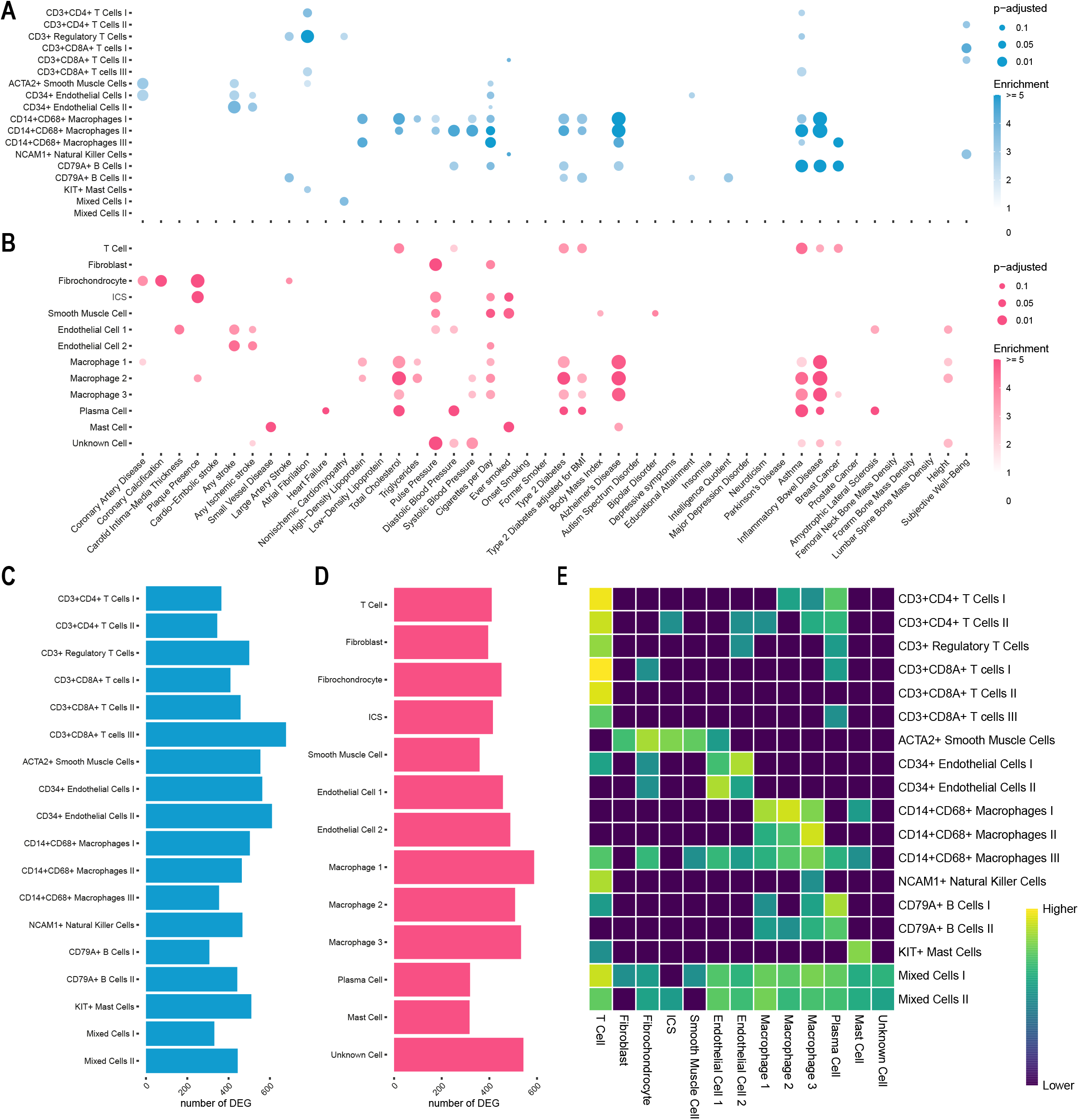
Gene score results for 46 GWAS in atherosclerotic plaque. **A**) Plot representing overlaps of GWAS candidate genes on cardiovascular disease and cardiometabolic traits with genes specifically expressed in cell populations from human atherosclerotic plaque from AtheroExpress biobank study. Colour intensity indicates enrichment over random data and size indicates p value (Wilcoxon rank sum test). **B**) Equivalent to **A**, but cell populations from atherosclerotic plaques from Pan *et al*.. **C**, **D**) Bar graph representing the numbers of DEGs per cell population. **E**) Heatmap showing similarity between the two different plaque datasets as determined with SingleR (log2 transformed data).

**Supplemental Figure 4.**
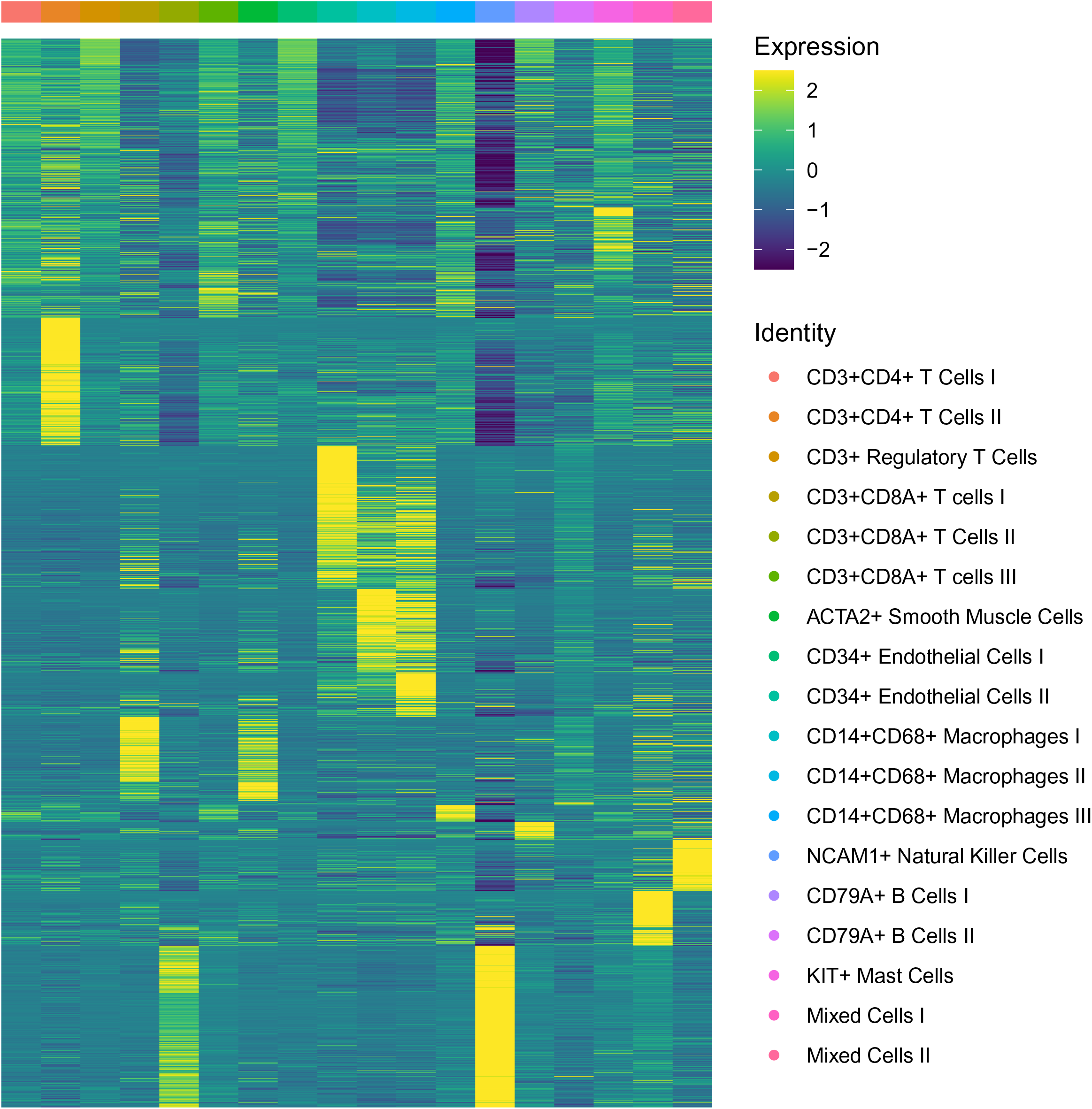
Heatmap with the DEG distribution of carotid atherosclerotic plaque. Average expression of cell-specific DEGs in cell populations from human atherosclerotic plaque.

**Supplemental Figure 5.**
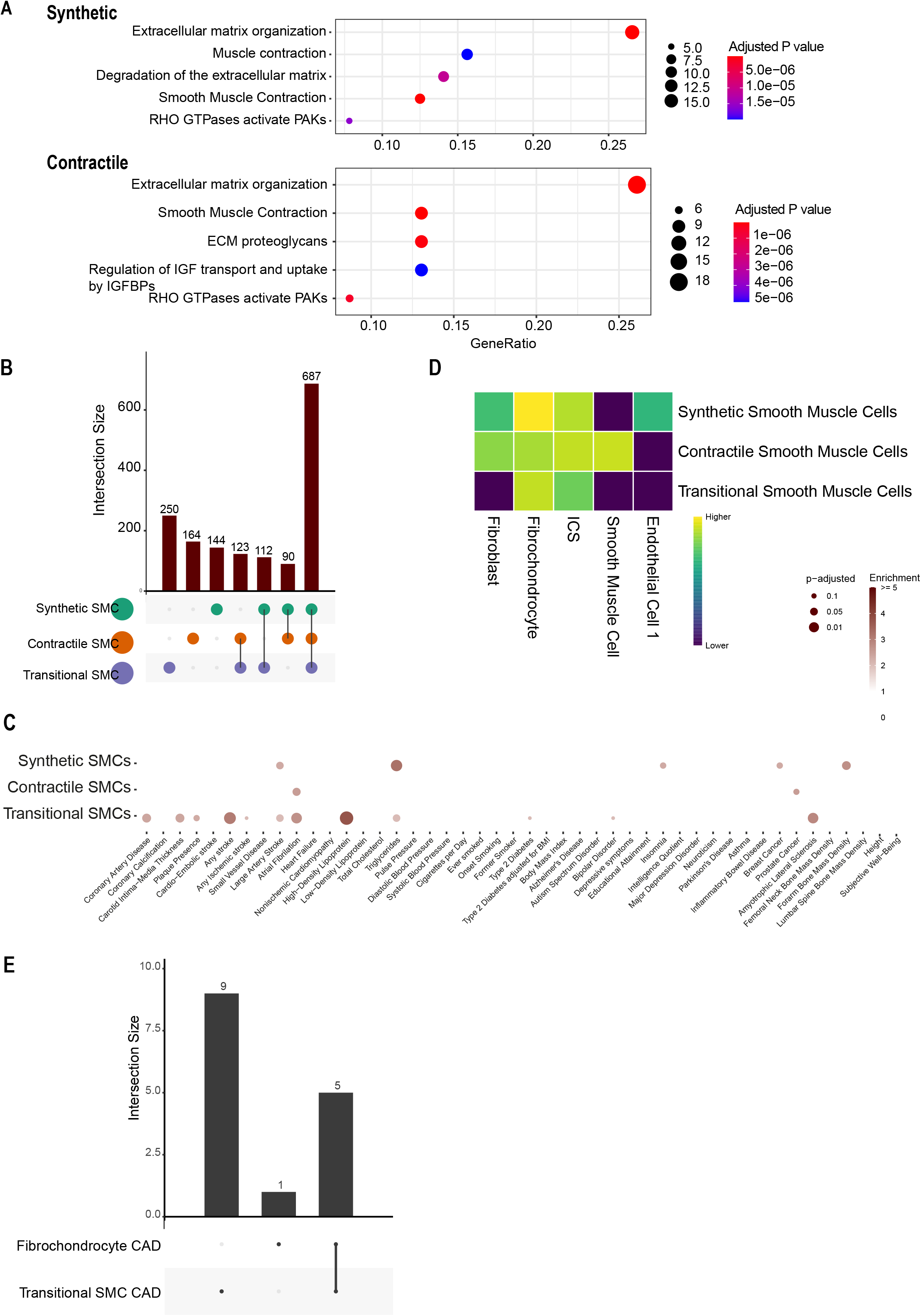
Smooth Muscle Cell population can be divided into three sub-populations. **A**) Pathway analysis of DEGs specific for synthetic and contractile SMCs. **B**) Plot depicting the number of unique and intersecting DEGs per sub-population. **C**) Plot representing overlaps of GWAS candidate genes on cardiovascular disease and cardiometabolic traits with genes specifically expressed in the subset of SMCs from human atherosclerotic plaque. Colour intensity indicates enrichment over random data and size indicates p value (Wilcoxon rank sum test). **D**) Heatmap showing similarity between SMC sub-populations from Athero-Express and Pan *et al.* as determined with SingleR (log2 transformed data). **E**) Plot depicting the number of unique and intersecting candidate genes from CAD in Fibrochondrocytes and Transitional SMCs.

**Supplemental Figure 6.**
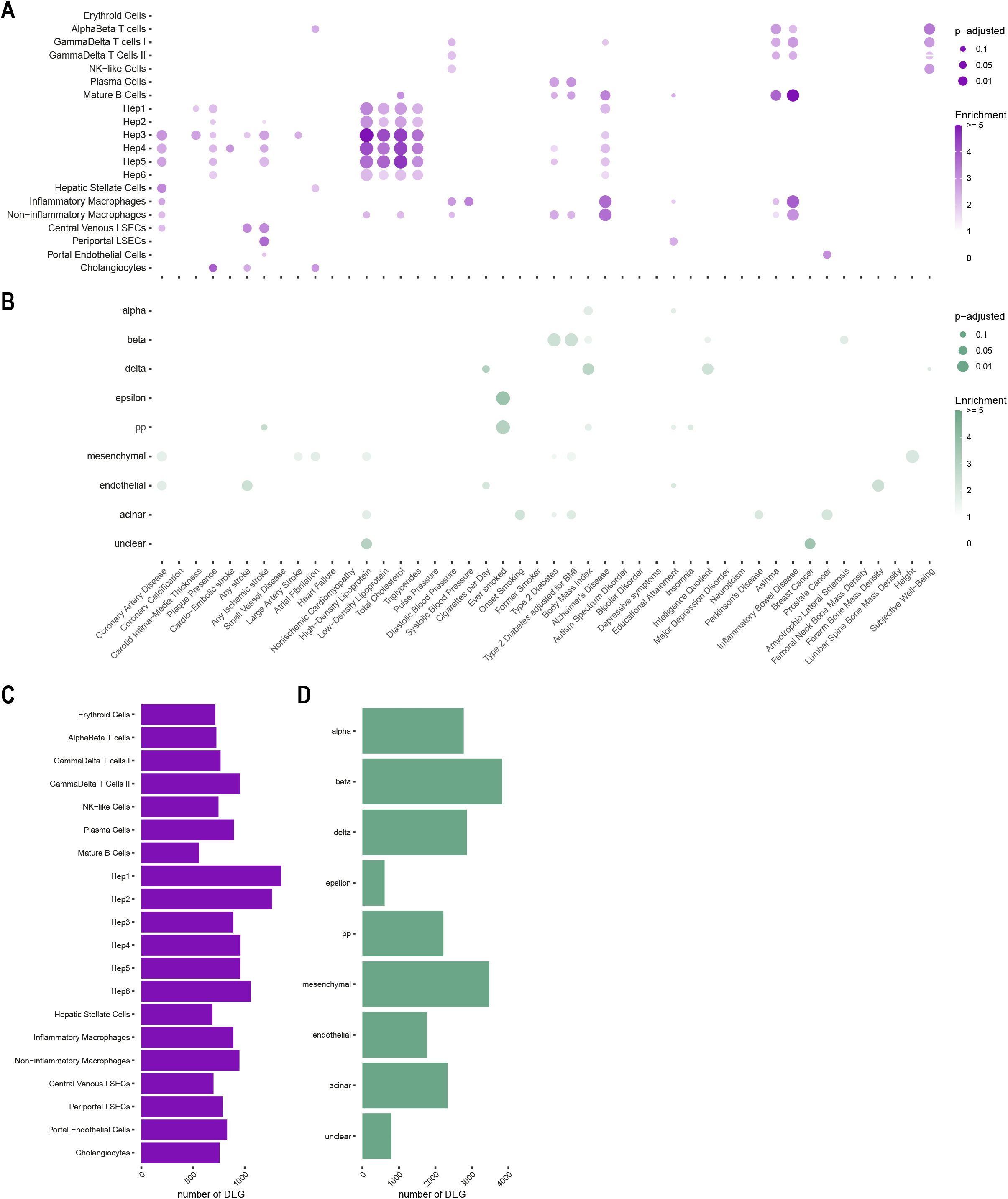
Gene score results for 46 GWAS in liver and Langerhans islets. **A**) Plot representing overlaps of GWAS candidate genes on cardiovascular disease and cardiometabolic traits with genes specifically expressed in cell populations from human liver from MacParland *et al*.. Colour intensity indicates enrichment over random data and size indicates p value (Wilcoxon rank sum test). **B**) Equivalent to **A**, but in cell populations from Langerhans islets from Muraro *et al*.. **C**, **D**) Bar graph representing the numbers of DEGs per cell population.

