## Supplemental Table 1 for "Intersecting single-cell transcriptomics and genome-wide association studies identifies crucial cell populations and candidate genes for atherosclerosis"

**Supplemental Table 1. List of included GWAS.**

| | Trait or disease | Abbreviation | Category | Sample size | Consortium | PMID | Reference | Journal | Ancestry |
|---|---|---|---|---|---|---|---|---|---|
| 1 | Coronary Artery Disease | CAD | Atherosclerotic disease | 154654 | CARDIoGRAMplusC4D+UKBB | 28714975 | Nelson et. al, 2017[3] | nature genetics | European |
| 2 | Coronary Artery Calcification | CAC | Atherosclerotic disease | 15523 | | 23561647 | Van Setten et al. 2013[5] | Atherosclerosis | European |
| 3 | Carotid Intima-Media Thickness | cIMT | Atherosclerotic disease | 71128 | CHARGE | 30510157 | Franceschini et al. 2018[6] | nature communication | Multi ancestry |
| 4 | Plaque Presence | Plaque | Atherosclerotic disease | 48434 | CHARGE | 30510157 | Franceschini et al. 2018[6] | nature communication | Multi ancestry |
| 5 | Cardio-Embolic stroke | CES | Cardiometabolic traits | 521612 | MEGASTROKE | 29531354 | Malik et al., 2019[4] | nature genetics | European |

| | | | | | | | | | |
|---|---|---|---|---|---|---|---|---|---|
| 6 | Any stroke | AS | Cardiometabolic traits | 446696 | MEGASTROKE | 29531354 | Malik et al., 2019[4] | nature genetics | European |
| 7 | Any Ischemic stroke | IS | Cardiometabolic traits | 446696 | MEGASTROKE | 29531354 | Malik et al., 2019[4] | nature genetics | European |
| 8 | Small Vessel Disease | SVD | Cardiometabolic traits | 446696 | MEGASTROKE | 29531354 | Malik et al., 2019[4] | nature genetics | European |
| 9 | Large Artery Stroke | LAS | Cardiometabolic traits | 446696 | MEGASTROKE | 29531354 | Malik et al., 2019[4] | nature genetics | European |
| 10 | Atrial Fibrillation | AF | Cardiometabolic traits | 588190 | | 29892015 | Roselli et al., 2018[43] | nature genetics | Multi ancestry |
| 11 | Heart Failure | HF | Cardiometabolic traits | 488010 | | 30586722 | Aragam et al. 2018[44] | Circulation | European |
| 12 | Nonischemic Cardiomyopathy | NICM | Cardiometabolic traits | 488010 | | 30586722 | Aragam et al. 2018[44] | Circulation | European |

| | | | | | | | | | |
|---|---|---|---|---|---|---|---|---|---|
| 13 | High-Density Lipoprotein | HDL | Cardiometabolic traits | 1888577 | GLGC | 24097068 | Global lipids Genetiscs Consortium , 2013[30] | nature genetics | European |
| 14 | Low-Density Lipoprotein | LDL | Cardiometabolic traits | 1888577 | GLGC | 24097068 | Global lipids Genetiscs Consortium , 2013[30] | nature genetics | European |
| 15 | Total Cholesterol | TC | Cardiometabolic traits | 1888577 | GLGC | 24097068 | Global lipids Genetiscs Consortium , 2013[30] | nature genetics | European |
| 16 | Triglycerides | TG | Cardiometabolic traits | 1888577 | GLGC | 24097068 | Global lipids Genetiscs Consortium , 2013[30] | nature genetics | European |
| 17 | Pulse Pressure | PP | Cardiometabolic traits | 1050906 | | 30224653 | Evangelou et al., 2018[45] | nature genetics | European |

| | | | | | | | | | |
|---|---|---|---|---|---|---|---|---|---|
| 18 | Diastolic Blood Pressure | DBP | Cardiometabolic traits | 1050906 | | 30224653 | Evangelou et al., 2018[45] | nature genetics | European |
| 19 | Systolic Blood Pressure | SBP | Cardiometabolic traits | 1050906 | | 30224653 | Evangelou et al,. 2018[45] | nature genetics | European |
| 20 | Cigarettes per Day | CpD | Cardiometabolic traits | 74035 | TAG | 20418890 | The Tobacco and Genetics Consortium, 2010[46] | nature genetics | European |
| 21 | Ever smoked | EvrSmk | Cardiometabolic traits | 74035 | TAG | 20418890 | The Tobacco and Genetics Consortium, 2010[46] | nature genetics | European |
| 22 | Onset Smoking | logOnset | Cardiometabolic traits | 74035 | TAG | 20418890 | The Tobacco and Genetics Consortium, 2010[46] | nature genetics | European |

| | | | | | | | | | |
|---|---|---|---|---|---|---|---|---|---|
| 23 | Former Smoker | FrmrSmk | Cardiometabolic traits | 74035 | TAG | 20418890 | The Tobacco and Genetics Consortium, 2010[46] | nature genetics | European |
| 24 | Type 2 Diabetes | T2D | Cardiometabolic traits | 898130 | DIAGRAM | 30297969 | Mahajan et al., 2016[47] | nature genetics | European |
| 25 | Type 2 Diabetes adjusted for BMI | T2DadjBMI | Cardiometabolic traits | 898130 | DIAGRAM | 30297969 | Mahajan et al., 2016[47] | nature genetics | European |
| 26 | Body Mass Index | BMI | Cardiometabolic traits | 693529 | GIANT | 30124842 | Yengo et al., 2018[48] | Human Molecular Genetics | European |
| 27 | Alzheimer's Disease | AD | Other | 455258 | | 30617256 | Jansen et al., 2019[49] | nature genetics | European |
| 28 | Autism Spectrum Disorder | ASD | Other | 46350 | PGC | 30804558 | Grove et al., 2019[50] | nature genetics | European |

| | | | | | | | | | |
|---|---|---|---|---|---|---|---|---|---|
| 29 | Bipolar Disorder | BIP | Other | 198882 | PGC | 31043756 | Stahl et al., 2019[51] | nature genetics | European |
| 30 | Depressive symptoms | DS | Other | 298420 | | 27089181 | Okbay et al., 2016[52] | nature genetics | European |
| 31 | Educational Attainment | EA | Other | 293723 | | 27225129 | Okbay et al., 2016[53] | nature genetics | European |
| 32 | Insomnia | Insomnia | Other | 386533 | | 30804565 | Jansen et al., 2018[49] | nature genetics | European |
| 33 | Intelligence Quotient | IQ | Other | 269867 | | 29942086 | Savage et al., 2018[54] | nature genetics | European |
| 34 | Major Depression Disorder | MDD | Other | 480359 | PGC | 29700475 | Wray et al., 2018[55] | nature genetics | European |
| 35 | Neuroticism | Neuroticism | Other | 298420 | | 27089181 | Okbay et al., 2016[52] | nature genetics | European |

| | | | | | | | | | |
|---|---|---|---|---|---|---|---|---|---|
| 36 | Parkinson's Disease | PD | Other | 1456306 | | 31701892 | Nalls et al.,2019[56] | The Lancet. Neurology | European |
| 37 | Asthma | Asthma | Other | 142486 | | 29273806 | Demenais et al., 2018[57] | nature genetics | Multi ancestry |
| 38 | Inflammatory Bowel Disease | IBD | Other | 86640 | | 26192919 | Liu et al., 2015[58] | nature genetics | Multi ancestry |
| 39 | Breast Cancer | BC | Other | 256123 | | 29059683 | Michailidou et al., 2017[59] | nature | Multi ancestry |
| 40 | Prostate Cancer | PrCa | Other | 140306 | | 29892016 | Schumacher et al., 2018[60] | nature genetics | European |
| 41 | Amyotrophic Lateral Sclerosis | ALS | Other | 43259 | | 27455348 | van Rheenen et al., 2016[61] | nature genetics | European |
| 42 | Femoral Neck Bone Mass Density | FNBMD | Other | 53236 | GEFOS | 26367794 | Zheng et al., 2015[62] | nature | European |

| | | | | | | | | | |
|---|---|---|---|---|---|---|---|---|---|
| 43 | Forarm Bone Mass Density | FABMD | Other | 53236 | GEFOS | 26367794 | Zheng et al., 2015[62] | nature | European |
| 44 | Lumbar Spine Bone Mass Density | LSBMD | Other | 53236 | GEFOS | 26367794 | Zheng et al., 2015[62] | nature | European |
| 45 | Height | Height | Other | 693529 | GIANT | 30124842 | Yengo et al., 2018[48] | Human Molecular Genetics | European |
| 46 | Subjective Well-Being | SWB | Other | 298420 | | 27089181 | Okbay et al., 2016[52] | nature genetics | European |
