## Supplemental Table 4 for "Intersecting single-cell transcriptomics and genome-wide association studies identifies crucial cell populations and candidate genes for atherosclerosis"

**Supplemental Table 4. scRNA-seq datasets.** n samples: number of patients used; n cells: number of cells per scRNA-seq dataset.

| | **reference** | **n samples** | **n cells** |
|---|---|---|---|
| **Atherosclerotic plaque** | | 38 | 6,191 |
| **Atherosclerotic plaque** | Pan *et al.,* 2020[23] | 3 | 8,867 |
| **Liver** | MacParland *et al.,* 2018[28] | 5 | 8,444 |
| **Langerhans islets** | Muraro *et al.,* 2016[29] | 5 | 2,292 |
