## Supplemental Table 5 for "Intersecting single-cell transcriptomics and genome-wide association studies identifies crucial cell populations and candidate genes for atherosclerosis"

**Supplemental Table 5. Functional assessment of smooth muscle cells.** Adjusted p-value (Benjamini Hochberg) and R of the correlation of gene expression of KANK2, SKI and EDNRA in (non)-proliferative cells in vascular SMCs. AUC: area under the curve; Pi: inorganic phosphate.

| | | **KANK2** | | **SKI** | | **EDNRA** | |
|---|---|---|---|---|---|---|---|
| | | p-adj | R | p-adj | R | p-adj | R |
| non-proliverative cells | Difference in AUC (Migration) | 0.227368421052632 | 0.15 | 0.425806451612903 | 0.1 | 0.430769230769231 | 0.096 |
| | Relative proliferation Control | 0.0617142857142857 | 0.23 | 0.228 | 0.15 | 0.465 | 0.087 |
| | Relative proliferation TFGβ1 | 0.0685714285714286 | 0.22 | 0.413793103448276 | 0.2 | 0.430769230769231 | 0.097 |
| | Relative proliferation PDGFBB | 0.0129 | 0.3 | 0.19125 | 0.17 | 0.413793103448276 | 0.11 |
| | Relative proliferation IL1β | 0.28695652173913 | 0.14 | 0.0685714285714286 | 0.22 | 0.883018867924528 | -0.024 |
| | Proliferation response TFGβ1 | 0.740425531914894 | -0.047 | 0.482926829268293 | -0.081 | 0.96 | 0.0056 |
| | Proliferation response PDGFBB | 0.430769230769231 | 0.093 | 0.96 | 0.0038 | 0.922222222222222 | 0.018 |
| | Proliferation response IL1β | 0.0617142857142857 | -0.22 | 0.3 | 0.13 | 0.038 | -0.26 |
| | Calcification (Pi) | 0.00582 | -0.28 | 0.796153846153846 | -0.035 | 0.197647058823529 | -0.17 |
| | Calcification Osteo | 0.0685714285714286 | -0.21 | 0.504545454545455 | 0.079 | 0.0617142857142857 | -0.23 |
| proliferative cells | Difference in AUC (Migration) | 0.0685714285714286 | 0.2 | 0.746938775510204 | 0.043 | 0.0685714285714286 | 0.21 |
| | Relative proliferation Control | 0.076 | 0.2 | 0.430769230769231 | 0.28 | 0.430769230769231 | 0.097 |
| | Relative proliferation TFGβ1 | 0.22 | 0.16 | 0.951724137931035 | -0.0082 | 0.430769230769231 | 0.098 |
| | Relative proliferation PDGFBB | 0.0617142857142857 | 0.23 | 0.796153846153846 | 0.037 | 0.485714285714286 | 0.082 |
| | Relative proliferation IL1β | 0.28695652173913 | 0.14 | 0.413793103448276 | 0.11 | 0.42 | 0.11 |
| | Proliferation response TFGβ1 | 0.430769230769231 | -0.095 | 0.0685714285714286 | -.021 | 0.730434782608696 | -0.049 |
| | Proliferation response PDGFBB | 0.746938775510204 | 0.044 | 0.430769230769231 | -0.091 | 0.413793103448276 | -0.11 |
| | Proliferation response IL1β | 0.312 | -0.13 | 0.6 | 0.063 | 0.951724137931035 | -0.0094 |
| | Calcification (Pi) | 0.28695652173913 | -0.14 | 0.932142857142857 | 0.87 | 0.796153846153846 | 0.036 |
| | Calcification Osteo | 0.488372093023256 | -0.082 | 0.0685714285714286 | 0.21 | 0.932142857142857 | 0.015 |
